# Corticospinal suppression underlying intact movement preparation fades in late Parkinson’s disease

**DOI:** 10.1101/2022.02.03.22269055

**Authors:** Emmanuelle Wilhelm, Caroline Quoilin, Gerard Derosiere, Susana Paço, Anne Jeanjean, Julie Duque

**Author notes:** Passed away on 15^th^ January 2020 (Anne Jeanjean). <u>Correspondence to:</u> Emmanuelle WILHELM, Institute of Neuroscience CoActions Lab, Université catholique de Louvain, 53, Avenue Mounier, COSY- B1.53.04, 1200 Brussels - BELGIUM.

## Abstract

In Parkinson’s disease, neurophysiological abnormalities within the primary motor cortex have been shown to contribute to cardinal symptoms such as bradykinesia, but the exact modalities are still uncertain. Here, we propose that such impairment could involve alterations of mechanisms shaping motor activity specifically during voluntary movement preparation. Indeed, several past studies in healthy populations have suggested that a strong suppression of corticospinal excitability called “preparatory suppression” – propels movement execution by increasing motor neural gain. Thus, we hypothesized that a gradual alteration to this mechanism may contribute to progressive motor slowness in Parkinson’s disease. We also predicted a benefit of dopamine medication in restoring correct motor neural activity during action preparation.

To test these hypotheses, we investigated preparatory suppression on two consecutive days in 29 Parkinson’s disease patients (ON and OFF medication) and 29 matched healthy controls. Single-pulse transcranial magnetic stimulation was applied over both primary motor cortices, eliciting concurrent motor-evoked potentials in the two hands, while subjects were either at rest or prepared a left- or right-hand response in an instructed-delay choice reaction time task. Preparatory suppression was assessed by expressing the amplitude of motor potentials evoked during movement preparation relative to those obtained at rest. These neurophysiological measures were cross-analysed with task behaviour and clinical data.

Contrary to healthy controls, Parkinson’s disease patients showed a lack of preparatory suppression, which appeared to depend on disease progression, but not on dopamine medication. Indeed, in relatively early disease stages, patients still exhibited partial preparatory suppression, while in later stages, they lacked it completely and even exhibited a tendency for corticospinal facilitation in the hand selected for movement execution. As expected, patients also showed increasing motor handicap with disease progression as well as a decreased movement velocity during the task, but such findings did not directly correlate with levels of preparatory suppression in our cohort. While dopamine medication had no effect on the latter, it did however globally reduce raw corticospinal excitability in the dominant hand.

Taken together, our results are in line with the idea that a lack of corticospinal suppression during movement preparation in Parkinson’s disease slows down response execution and illustrate the importance of considering disease stages in such investigations; they also suggest differential roles of dopamine in shaping corticospinal output in those patients. Our findings thus support the use of task-related functional markers such as preparatory suppression in future studies on motor impairment in Parkinson’s disease.

## Introduction

Parkinson’s disease is a progressive neurodegenerative disorder, defined as the presence of bradykinesia combined with rest tremor, rigidity or both, according to current diagnostic criteria [1, 2]. Bradykinesia thus counts among the cardinal motor symptoms and is characterized, among others, by slowness and reduced amplitude of voluntary movements [3]. Its pathophysiology is strongly linked to the nigrostriatal neurodegeneration pathognomonic of Parkinson’s disease, which causes a significant dopamine depletion in the striatum [4, 5]. The resulting dysfunction in cortico-basal ganglia loops leads to a decreased output from the thalamus onto the primary motor cortex (M1) [6, 7] and a disruption of the preparatory and executive phases of voluntary movements, clinically translating into deficits such as bradykinesia [3, 8–11]. Interestingly, major research advances, mostly over the last decade, have shown that alterations to the activity within M1 and to deriving corticospinal motor projections directly contribute to the pathophysiology of Parkinson’s disease [12–17]. How exactly these intrinsic motor neural abnormalities are linked to movement deficits such as bradykinesia is, however, still a matter of debate [3]. Here, we propose that bradykinesia in Parkinson’s disease could be linked to pathological changes in patterns of corticospinal excitability supporting voluntary movement preparation, typically observed in healthy subjects.

Both motor cortical and output activity strongly depend on the balance between excitatory and inhibitory influences which, in humans, can be explored non-invasively with single-pulse transcranial magnetic stimulation (TMS) over M1, eliciting motor evoked potentials (MEPs) in targeted muscles [18–20]. In healthy participants, studies using TMS over M1 have repeatedly observed a transient drop in MEP amplitudes during movement preparation, suggesting a paradoxical suppression of excitability within the corticospinal tract preceding the execution of an action [21–23]. Even if this so-called “preparatory suppression” of the motor system is highly reproducible, its nature has not been completely elucidated yet [22, 24–26].

For a long time, preparatory suppression was solely linked to behavioural inhibition, offering a means to prevent premature actions during motor planning, as supported by findings in clinical populations characterized by impulsive behaviours [23, 27–30]. More recently, however, a different hypothesis has emerged, according to which preparatory suppression could also facilitate movement release and speed [22, 24, 31] in a “gain modulation” fashion [32–34]. Following the latter idea, greater inhibition of the motor system could enhance the sensitivity of the selected movement representation to excitatory inputs [35], thus increasing the signal-to-noise ratio and favouring rapid response execution [36]. Such a mechanism could possibly overlap with the effect of the behavioural inhibition process, both contributing to MEP measures of preparatory suppression [26]. In line with this idea, a few recent studies in healthy human participants revealed that the strength of preparatory suppression correlates with the speed of motor responses in various reaction time (RT) tasks: the stronger the suppression of motor activity during movement preparation, the faster the response execution [22, 31]. Dopamine appears as a likely source for this mechanism given its role in activating cortical interneurons and in increasing the signal-to-noise ratio in cortical regions [37]; this neuromodulator has also been shown to enhance the gain of cells, both in the striatum [38, 39] and in the motor output pathway [40].

In Parkinson’s disease, neurophysiological studies using TMS have already allowed to uncover abnormal changes within the motor system, typically showing attenuated levels of M1 inhibition accompanied by an increase in corticospinal output [8, 14–16, 41–43]; yet, most of these works acquired measures at rest or during simple RT tasks, without probing corticospinal excitability specifically during movement preparation. Interestingly though, previous studies indicate that the abnormal M1 and corticospinal excitability at rest in Parkinson’s disease patients can be normalized by dopaminergic medication [3, 15, 44], in parallel to the well-known alleviating effect of this treatment on motor parameters, such as movement speed and amplitude [45].

Based on the idea that aspects of preparatory suppression facilitate the release and speed of selected movements, and based on the recurrent findings of increased corticospinal output in Parkinson’s disease, here we tested the hypothesis that bradykinesia is linked to a lack of corticospinal suppression during movement preparation. Furthermore, we postulated that dopamine intake would reduce motor slowness in patients at least in part by restoring preparatory suppression. Finally, given that dopaminergic neurodegeneration increases as a function of disease duration [46], we expected deficits in preparatory suppression in Parkinson’s disease to worsen with disease progression [47], thus contributing to increasing bradykinesia [11] in a dopamine-dependent fashion. To address these hypotheses, we tested 29 Parkinson’s disease patients matched with an equal number of healthy control (HC) subjects, with each participant undergoing two testing sessions on two consecutive days. In each session, we used single-pulse TMS over M1 to elicit MEPs in participants performing an instructed-delay choice RT task, which allowed us to measure corticospinal excitability during movement preparation. In order to assess the possible effect of dopaminergic treatment on our measures, in each session, patients were tested either OFF or ON their usual dopamine replacement therapy (DRT), in a randomized order; both sessions were identical in healthy subjects. Motor impairment was quantified in both sessions and for all patients using the motor assessment of the MDS Unified Parkinson’s disease Rating Scale (MDS-UPDRS, part III). All data were analysed using fully automatic procedures to avoid possible confirmation bias.

## Materials and methods

### Participants

In total, we recruited 29 patients with idiopathic Parkinson’s disease 13 females, mean age 64.7 ± 10.5 years) and 29 HC subjects, matched for age, gender and years of education (mean age 62.9 ± 9.5 years). All patients were diagnosed by a certified neurologist with specialty training in movement disorders, based on the clinical diagnostic criteria validated by the Movement Disorder Society [2, 48, 49] Parkinson’s disease patients were recruited through the Adult Neurology department of the Saint-Luc University Hospital (Brussels, Belgium), whereas HC subjects were recruited through hung-out flyers and social media groups. All patients were treated by DRT and were physically independent enough to come to the laboratory on their own (Hoehn & Yahr stage ≤ 3, with and without dopamine medication) [50]. None of them had impulse control disorders or dopamine dysregulation syndrome [51], in order to preclude a lack of preparatory suppression due to a deficit in behavioural inhibition [27]. Besides, we did not observe apparent dyskinesias during the sessions; tremor, if present, was sufficiently mild not to interfere with the experiments. The clinical details of patients are summarized in Table 1.

**Table 1.** Details of demographic and clinical data in Parkinson’s disease patients.

| Gender | Handedness | Years since diagnosis | Hoehn & Yahr | MDS-UPDRS-III, OFF-DOPA | MDS-UPDRS-III, ON-DOPA | LEDD (mg) | Dominant side of PD | MoCA | BDI-II | UPPS |
| --- | --- | --- | --- | --- | --- | --- | --- | --- | --- | --- |
| M | R | 1 | 2 | 24 | 19 | 400 | L | 25 | 13 | 107 |
| F | R | 1 | 2 | 39 | 34 | 200 | L | 27 | 14 | 102 |
| F | R | 2 | 2 | 21 | 16 | 450 | R | 27 | 3 | 81 |
| M | L | 2 | 2 | 23 | 23 | 126 | R | 28 | 3 | 81 |
| M | R | 3 | 2 | 12 | 6 | 790 | R | 28 | 6 | 57 |
| M | R | 3 | 2 | 25 | 15 | 610 | R | 27 | 3 | 111 |
| M | R | 3 | 2 | 15 | 16 | 600 | R | 25 | 13 | 100 |
| F | R | 3 | 2 | 17 | 16 | 330 | L | 26 | 8 | 114 |
| M | R | 4 | 2 | 18 | 12 | 652 | L | 27 | 9 | 63 |
| M | R | 4 | 2 | 44 | 46 | 400 | L | 26 | 12 | 72 |
| M | R | 4 | 2 | 28 | 24 | 1895 | R | 29 | 14 | 81 |
| F | R | 5 | 2 | 9 | 3 | 760 | R | 28 | 5 | 85 |
| M | R | 5 | 2 | 29 | 17 | 810 | L | 29 | NA | 89 |
| M | R | 5 | 2 | 28 | 19 | 985 | L | 28 | 5 | 65 |
| F | R | 5 | 2 | 28 | 17 | 760 | L | 30 | 21 | 103 |
| F | R | 5 | 2 | 29 | 15 | 775 | R | 26 | 15 | 85 |
| F | R | 6 | 3 | 38 | 29 | 864 | R | 29 | 13 | 80 |
| M | L | 8 | 3 | 36 | 27 | 953 | L | 27 | 24 | 104 |
| M | L | 8 | 2 | 32 | 26 | 805 | R | 29 | 21 | 103 |
| F | R | 8 | 3 | 51 | 37 | 650 | L | 29 | 23 | 95 |
| F | R | 9 | 2 | 38 | 17 | 771 | R | 27 | 19 | 85 |
| F | L | 9 | 2 | 25 | 16 | 540 | R | 26 | 7 | 70 |
| F | R | 10 | 2 | 37 | 25 | 610 | R | 28 | 11 | 115 |
| M | R | 11 | 3 | 35 | 26 | 875 | L | 29 | 8 | 95 |
| M | R | 11 | 2 | 40 | 33 | 460 | R | 27 | 9 | 86 |
| M | R | 12 | 2 | 44 | 27 | 890 | L | 26 | NA | NA |
| M | R | 13 | 2 | 42 | 34 | 938 | L | 29 | 12 | 59 |
| F | R | 15 | 2 | 33 | 27 | 745 | L | 30 | 13 | 80 |
| F | R | 16 | 3 | 37 | 28 | 680 | L | 26 | 15 | 91 |

PD = Parkinson's disease; LEDD = Levodopa Equivalent Daily Dose; ON-DOPA = session performed one hour after the intake of dopamine replacement therapy; OFF-DOPA = session performed after overnight withdrawal of dopamine replacement therapy (min. 12 hours withdrawal for Levodopa and min. 24h withdrawal for dopamine agonists (and other Parkinson's disease medication)); MoCA = Montreal Cognitive Assessment; BDI-II = Beck Depression Inventory, Second Edition; UPPS = UPPS Impulsive Behaviour Scale; NA = not available. Note that patients are ranged according to the years since diagnosis; the horizontal line after six years since diagnosis indicates the cut-off for the subgroup split.

Based on a condensed version of the Edinburgh Handedness Inventory [52], 25 patients and all controls were right-handed. Exclusion criteria for both groups comprised: (1) presence of severe cognitive decline (based on the Montreal Cognitive Assessment) [53, 54], (2) MRI-incompatible metal device in the body, (3) history of major psychiatric or neurological disorder (other than Parkinson’s disease for the patient group), (4) history of substance use disorder (except nicotine), (5) untreated or unstable medical conditions that could interfere with cognitive functioning.

Participants were naïve to the purpose of the study and gave written informed consent, following a protocol approved by the Biomedical Ethics Committee of the Saint-Luc University Hospital (*UCLouvain*, Brussels, Belgium; 2018/27AVR/194 - B403201836769), in accordance with the Declaration of Helsinki. A financial compensation was provided to all participants, in accordance with the ethical protocol.

## Experimental procedure

Data acquisition was performed at the Institute of Neuroscience of *UCLouvain* (Brussels, Belgium). Every participant had to undergo two separate testing sessions on two consecutive days, at the same hour in the morning. Parkinson’s disease patients performed one of the sessions one hour after the intake of their first dose of daily DRT (hereafter referred to as the “ON-DOPA” condition), which, allowed to test them at their expected peak of Levodopa [55]. The other session always occurred after an overnight withdrawal of their medication (“OFF-DOPA” condition); in that case, we made sure that the last intake of Levodopa and dopamine agonists (and any other Parkinson’s disease medication) happened at least 12 and 24 h before the beginning of the testing, respectively [56, 57]. The order of the sessions was randomized, with 15 patients starting the experiment with the OFF-DOPA session and 14 with the ON-DOPA session. In order to evaluate their level of motor impairment according to the presence or absence of dopaminergic medication at the moment of the testing, patients underwent the MDS-UPDRS part III at the beginning of each session, with the same trained physician [58]. Matched controls performed two identical sessions on two consecutive days, at the same time of the day as the patients. For each HC subject, we randomly turned one of their sessions into a fictive OFF-DOPA session while turning the other one into a fictive ON-DOPA session, in a completely randomised way (both sessions were identical in HCs): in accordance with the patients, 15/29 control subjects were attributed a fictive OFF-DOPA session on their first day of testing. All participants filled in the French version of two self-evaluation questionnaires: the Beck Depression Inventory Second Edition (BDI-II) [59–61] and the UPPS Impulsive Behaviour Scale [62, 63] (measuring trait impulsivity), both validated for the use in Parkinson’s disease [64–66].

### Response device and rolling ball task

After having undergone all pre-cited evaluations and questionnaires, for the main part of the experiment, participants were installed in a quiet and dimly-lit room, in front of a computer screen positioned about 60 cm in front of them, with both forearms resting on a table in a semi-flexed position. Participants performed an instructed-delay choice RT task involving left or right index finger responses, which was implemented by means of Matlab 7.5 (The Mathworks, Natick, MA, USA) using the Psychophysics Toolbox extensions [67, 68]. The subjects’ hands were placed palms down on a homemade response device (one for each hand) allowing to detect any horizontal movement of the index fingers [29, 30, 69]. Each response device was composed of one horizontal outer and one vertical inner metal plate (Fig.1.A). At rest, subjects were asked to relax their index fingers in a reference position on the horizontal plate. The device registered a response when participants performed a brisk abduction movement of the left or right index finger and touched the internal vertical plate; the arrival time (AT) was defined as the time needed to reach the inner metal contact. Subjects always had to go back to the outer reference position before providing the next response.

**Figure 1.**
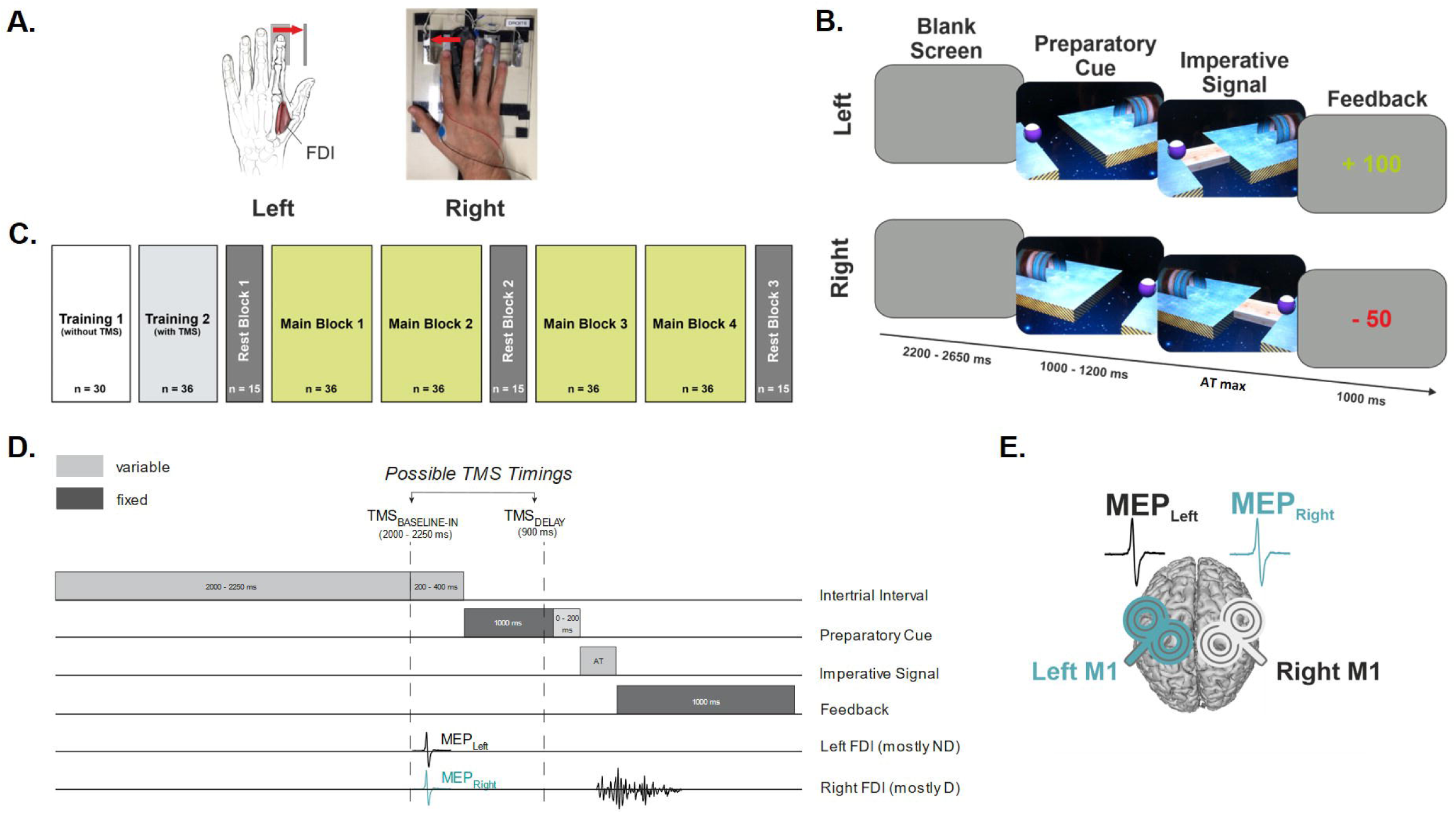
Experimental procedure. **A. Response device and EMG recording**. A response device was positioned under each hand (left = graphic representation; right = photographic representation). Each device was composed of a pair of metal edges fixed on a plastic support; the distance separating the centre of the outer horizontal from the inner vertical plate was 2 cm. This device detected responses by registering the abduction movements from the outer to the inner metal edge of the left and right index fingers. It allowed for a very precise measure of the arrival time (AT) (precision = 1 ms) while controlling for the initial position of the fingers. EMG activity was recorded from surface electrodes placed on the first dorsal interosseous (FDI) in both hands. (Note that, for the purpose of a separate study, electrodes were also placed on two other muscles, but are not shown on the figure for more clarity). **B. Rolling ball task**. Participants performed an instructed-delay choice reaction time task, in which they had to choose between a left or right index finger abduction (left and right example shown) according to the position of a preparatory cue (a ball). They were asked to withhold their response until the onset of an imperative signal (a bridge). Once the bridge appeared, they had to release their response as quickly as possible, with the correct finger, in order to make the ball roll over the bridge into the goal located on the other side of the gap. A feedback of the response appeared at the end of each trial, in green or red, following a correct or incorrect answer, respectively. **C. Experimental design**. After two training blocks, participants performed four main blocks of *n* = 36 trials each, during which motor-evoked potentials (MEPs) were elicited either at TMS_BASELINE-IN_ or TMS_DELAY_, in a random order. MEPs were also elicited in three rest blocks, outside the context of the task (TMS_BASELINE-OUT_; before main block 1, after main block 2 and after main block 4). **D. Time course of a trial**. Each trial began with the presentation of the preparatory cue displayed on the screen for a variable duration (1000—1200 ms), followed by the imperative signal which remained visible until a finger response was provided (the maximum time given to respond was individualised for each participant based on the training performance). Then, a feedback score based on the trial’s performance appeared for 1000 ms, which was followed by a blank screen lasting for a variable interval of 2200–2650 ms. TMS pulses could occur either during the inter-trial interval (between 2000 and 2250 ms after the onset of the blank screen; TMS_BASELINE-IN_) or during the delay period (900 ms after the onset of the preparatory cue; TMS_DELAY_). Since 93% of participants were right-handed, the right FDI corresponded to the dominant (D) effector in the majority of cases, whereas the left FDI mostly corresponded to the non-dominant (ND) one. This example shows a right finger movement response, which corresponded to the dominant side (RESP_D_) in the majority of participants. **E. TMS protocol**. Two small figure-of-eight coils were placed over both primary motor cortices for each participant, eliciting MEPs in the left and right hands with a 1 ms inter-pulse interval.

The task consisted in a virtual 3D “Rolling Ball” game previously used in several other studies [27, 29, 31, 69–72] (Fig.1.B). In this game, participants were instructed to choose between an index finger abduction with the left or right hand, corresponding mostly to the non-dominant (ND) or dominant (D) hand, respectively (in all healthy controls and 86% of Parkinson’s disease patients); the choice had to be made according to the position of a preparatory cue, which appeared on the computer screen in front of them. This cue, under the form of a ball, was presented either on the left or the right side of the screen, opposite to a goal and separated from it by a gap. The side of the ball indicated the side required for the forthcoming movement response. Importantly, participants had to prepare but withhold their abduction movement for a period lasting 1000-1200 ms, until an imperative signal appeared, which was a bridge closing the gap between the ball and the goal. The duration of this preparatory delay was purposely variable in order to decrease participants’ tendency to respond prematurely (i.e. before the imperative signal). Note that responses provided before the appearance of the imperative signal caused the ball to fall into the gap. To further prevent subjects from anticipating, we also included a few catch trials (6 % of trials) in which the ball was not followed by a bridge; participants were instructed not to respond on these trials and were penalized if they still did. As soon as the imperative signal appeared, participants had to provide a response as quickly as possible, with the correct index finger, hence making the ball roll over the bridge into the goal.

They had to respond within a maximum time that was individualized in each subject and corresponded to their median AT generated during the second training block (“Training 2”, preceding the start of the main experiment, see Fig.1.C) to which were added two standard deviations (SD). This maximum time corresponded to 463.2 ± 97.4 ms and 421.5 ± 82.0 ms in patients and controls, respectively, and did statistically differ between both groups as shown by an independent sample *t*-test (*t* = 2.4, *P* = 0.02). The imperative screen disappeared once a response was detected (or after the individualized maximum time) and a feedback was presented for 1000 ms. Finally, each trial ended with a blank screen, lasting 2200-2650 ms (inter-trial interval). The sequence and timing of events in each trial are shown in Figure 1.D. Following a correct response, participants received a feedback that consisted of a positive score inversely proportional to the trial’s AT (i.e., the faster the subjects, the higher the score), ranging from 1 to 100 and depicted in green at the centre of the screen (Vassiliadis et al., 2018). Note that when subjects succeeded not to respond in a catch trial, they received a positive feedback, whereas if they provided a response in the absence of the bridge, the ball fell into the gap and they received a negative score. Negative scores also occurred when subjects responded too early (i.e. before the imperative signal), too late (i.e. more than the individualized maximum time) or with the incorrect finger.

### TMS protocol

Monophasic pulses were delivered using a double-coil TMS method whereby pulses are delivered over the two M1 with a 1 ms inter-pulse interval, as used in many previous studies [25, 71, 73–75] (Fig.1.E). This technique allows eliciting near-simultaneous MEPs in both hands that are comparable to those elicited with single-coil TMS, regardless of the pulse order or the intensity of stimulation [72, 76]. In the present study, the first TMS pulse was always applied over the right M1, eliciting a first MEP in the left hand; a second MEP followed 1 ms later in the right hand [73]. The pulses were generated with two small figure-of-eight coils (wing internal diameter: 35 mm), as in most subjects it is not possible to simultaneously place two large coils over both M1. The coil stimulating the right M1 was connected to a Magstim 200 ^2^ magnetic stimulator (Magstim, Whitland, Dyfed, UK) and the coil stimulating the left M1 to a Magstim BiStim ^2^ magnetic stimulator. Both coils were placed tangentially over both M1 with the handle pointing backward and laterally at a 45° angle away from the midline, approximately perpendicular to the central sulcus, in order to induce a posterior-anterior current in the underlying neural cells [21, 25] (Fig.1.E). For each M1, we identified the optimal scalp position for eliciting contralateral MEPs in the First Dorsal Interosseus (FDI; task agonist), as well as in the Abductor Digiti Minimi (ADM) and Abductor Pollicis Brevis (APB) muscles. This multi-muscle hotspot was marked on a cap placed on the participant’s head to provide a reference mark throughout the experiment [29–31]. For each M1, the resting motor threshold was determined as the minimal TMS intensity required to evoke MEPs of minimum 50 µV peak-to-peak in each of the three muscles in at least 5 out of 10 consecutive trials [25]. Notably, MEPs elicited in the ADM and APB were obtained as part of another study; here we focus on the FDI, the prime-mover in the task. For each hemisphere, the intensity of TMS throughout the experiment was always set at 115% of the individual resting motor threshold [30], which allowed us to elicit reliable FDI MEPs bilaterally.

### Experimental design

The experiment started with two training blocks (Fig.1.C). The first block, Training 1, was composed of 30 rolling ball trials without TMS, during which participants discovered and practiced the task. The second block, Training 2, was composed of 36 rolling ball trials, but here TMS was applied in order to familiarize subjects with the sensation of pulses while performing the task. The latter block also served to calculate the median AT of each subject, which was used to 1) adapt the maximum time allowed to respond after the appearance of the imperative signal and 2) individualize the feedback scores on correct trials. Note that Training 1 had to be repeated once in some participants, in order to make sure that the indications of the virtual game were perfectly understood. Then, during the main phase of the experiment, all participants performed the task in four main blocks of 36 trials each; TMS was applied over both M1 in 30/36 trials. Each of those main blocks consisted of an equal proportion of left- and right-hand (corresponding to ND and D, respectively, in most subjects) response trials (i.e., 18 trials per hand condition, 1 of which was a catch trial). TMS pulses were delivered at one of two possible timings during the task (only one timing per trial, see Fig.1.D). First, in order to establish a baseline measure of corticospinal excitability, some trials involved a TMS pulse during the inter-trial interval (TMS_BASELINE-IN_; 10 trials/block), allowing to elicit MEPs within the context of the task but when the participants were at rest. Second, in other trials, a TMS pulse was delivered during the preparatory delay, between the appearance of the preparatory cue and the imperative signal (TMS_DELAY_; 20 trials/block); these pulses always occurred 900 ms after the onset of the preparatory cue, hence while subjects were withholding either a ND (RESP_ND_; 10 trials/block) or a D (RESP_D_; 10 trials/block) finger response. Based on previous studies, we assumed that MEPs in HCs would be strongly reduced at TMS_DELAY_, reflecting preparatory suppression when participants are withholding a motor response [19, 26]. On the other hand, as compared to control subjects, we expected Parkinson’s disease patients to show weaker levels of preparatory suppression in their OFF-DOPA session, which were possibly restored with DRT in the ON-DOPA session. The remaining trials within the task (6/36) did not involve any TMS pulse and were included to prevent participants from anticipating the occurrence of a pulse at TMS_DELAY_ whenever there had been no pulse at TMS_BASELINE-IN_. Finally, in addition to probing corticospinal excitability during the task, we also measured baseline MEPs outside this context, at complete rest in front of a blank screen (TMS_BASELINE-OUT_). These rest blocks each consisted of 15 TMS pulses (over both M1), applied every 4200-5100 ms, and were performed three times in each session: at the beginning, the middle and the end of the experiment (Fig.1.C). Note that for every participant, a short break was made between the second rest block and the third main block. Taken together, for each muscle and in each session, we thus obtained 45 MEPs at TMS_BASELINE-OUT_, 40 MEPs at TMS_BASELINE-IN_ and 80 MEPs at TMS_DELAY_ (i.e. 40 for each responding hand). The main blocks lasted ±6 minutes each, compared to ±2 minutes per rest block.

### EMG recording and data analysis

EMG activity was recorded from the FDI using surface electrodes (Ambu BlueSensor NF-50-K/12/EU, Neuroline, Medicotest, Oelstykke, Denmark) placed over the left- and right-hand muscles (Fig.1.A). EMG data was collected for 3000 ms on each trial, starting 200 ms before the TMS pulse; signals were monitored visually on a computer screen during the whole experiment. The raw EMG signals were amplified (gain of 1K), band-pass filtered on-line (10-500 Hz), notch-filtered (50 Hz) in case of line noise contamination (Digitimer D360, Hertfordshire, UK) and digitized at a sampling rate of 2 kHz (CED 1401-3 ADC12 and Signal 6 software, Cambridge Electronic Design Ltd) for off-line analysis. Data extraction and cleaning were performed using fully automated procedures in Signal 6 and R Studio (https://www.r-project.org/), respectively. The EMG signals were used to measure the peak-to-peak amplitude of FDI MEPs in the D (MEP_D_) and ND (MEP_ND_) hands as well as to extract the RT. The latter was defined as the moment after the imperative signal when the EMG activity started exceeding its mean by 3 SD and its root mean square value exceeded 0.2 [77]. The movement time (MT) was then obtained by subtracting the RT from the AT. Trials with any background EMG activity exceeding 3 SD above the mean of the root mean square in the 200 ms window preceding the TMS pulse were excluded from the analysis. This was done to prevent contamination of the MEP measurements by significant fluctuations in background EMG [29–31]. This is another reason why, in this study, we could not include Parkinson’s disease patients with significant rest tremor as their EMG background activity would have precluded us from retaining sufficient measurements for our MEP analyses. Trials in which subjects had made an error were also removed from the data set before further analysis. The remaining MEP_ND_ and MEP_D_ were classified according to the experimental condition in which they had been elicited (TMS_BASELINE-OUT_, TMS_BASELINE-IN_, TMS_DELAY_ with RESP_ND_ or RESP_D_). For each muscle and condition, we excluded trials with peak-to-peak MEP amplitudes larger or smaller than 3 SD around the mean. The number of trials left to asses MEPs in each muscle and condition after data cleaning are presented in the Supplementary Table 1.

## Statistical analysis

### Demographic and clinical data

Differences in age and BDI-II between Parkinson’s disease patients and HC subjects were evaluated using a Mann-Whitney U-test. Participants’ trait impulsivity was analysed by conducting a multivariate analysis of variance (MANOVA) on scores reported at the four subscales of the UPPS questionnaire.

In patients, the total scores on the MDS-UPDRS part III as well as the bradykinesia subscores (derived from the same clinical assessment) in the OFF- and ON-DOPA sessions were compared using a Wilcoxon test for each. Based on an inconsistent establishment of bradykinesia subscores in the literature [78–80], in the present study, we decided to include the items 3.2, 3.4, 3.5, 3.6, 3.7, 3.8 and 3.14 of the MDS-UPDRS part III, in order to englobe the widest set of motor characteristics related to this cardinal Parkinson’s disease symptom [3]. Finally, we checked how clinical scores at the MDS-UPDRS part III, OFF-DOPA, evolved with the number of years since the diagnosis of Parkinson’s disease, using ordinary least squares (OLS) regressions; this was done for both the total motor scores and the bradykinesia subscores. We focused on OFF-DOPA measures, as the absence of medication can be considered as the raw state of the disease and thus the true clinical reference condition. These regressions allowed us to check if the variable “years since diagnosis” in our patient cohort could be used as a marker of increased motor impairment, and thus as a probe of dopaminergic neurodegeneration. Whenever we performed OLS regressions, we first verified if there was no heteroscedasticity in the data, both visually as well as mathematically, using the Breusch-Pagan and the non-constant variance score test.

### Preparatory suppression in the rolling ball task

Next, in order to probe levels of preparatory suppression, we looked at MEPs acquired at TMS_DELAY_ and expressed them in percentage of MEPs elicited at TMS_BASELINE-IN_ in the corresponding hand. Group comparisons were then conducted using a four-way ANOVA with GROUP (Parkinson’s disease patients, HC subjects) as between-subject factor and with DRT (ON-DOPA, OFF-DOPA), RESP_SIDE_ (RESP_ND_, RESP_D_) and MEP_SIDE_ (MEP_ND_, MEP_D_) as within-subject factors. We checked the data for sphericity using the Mauchly test whenever we conducted a repeated-measures ANOVA. To assess the degree of preparatory suppression in each sub-condition, one-sample *t*-tests (Bonferroni-corrected) were carried out to compare these values to a reference value of 100 (i.e. to TMS_BASELINE-IN_). Next, we checked how these measures of corticospinal suppression evolved with the number of years since diagnosis of Parkinson’s disease, using OLS regressions.

### Reaction and movement times in the rolling ball task

In order to analyse the behaviour of participants during the rolling-ball task, we focused on trials in which the TMS pulses were applied during the inter-trial interval (TMS_BASELINE-IN_) or trials in which there was no TMS pulse at all, in order to avoid a possible disturbance effect of the TMS pulse; all those trials were pooled together. For the analyses of RT and MT, we conducted two separate three-way ANOVAs, with GROUP (Parkinson’s disease patients, HC subjects) as between-subject factor and with DRT (ON-DOPA, OFF-DOPA) and RESP_SIDE_ (RESP_ND_, RESP_D_) as within-subject factors. Here, we checked for a possible correlation between behavioural measures and levels of preparatory suppression, using again OLS regressions.

### Raw corticospinal excitability

In a final set of analyses, we investigated whether the two groups displayed differences in resting motor thresholds by means of a three-way ANOVA with GROUP (Parkinson’s disease patients, HC subjects) as between-subject factor and with DRT (ON-DOPA, OFF-DOPA) and HEMISPHERE_SIDE_ (HEMI_ND_, HEMI_D_) as within-subject factors. Then, we focused on measures of corticospinal excitability at rest by considering MEPs acquired at TMS_BASELINE-OUT_ (i.e. outside the main blocks) and at TMS_BASELINE-IN_ (i.e. within the main blocks). The raw amplitude of these MEPs (mV) was analysed using a four-way ANOVA, with GROUP (Parkinson’s disease patients, HC subjects) as the between-subject factor and with DRT (ON-DOPA, OFF-DOPA), BASELINE (TMS_BASELINE-OUT_, TMS_BASELINE-IN_) and MEP_SIDE_ (MEP_ND_, MEP_D_) as within-subject factors. Following these analyses, we also considered the raw amplitude of MEPs acquired at TMS_DELAY_, in addition to the analysis of preparatory suppression; these are presented in the Supplementary Fig.2.

For all ANOVAs, we ran post-hoc comparisons using Fisher’s Least Significant Difference method. Data is shown as mean ± SE and the statistical significance was set at P < 0.05. Analyses were carried out using Statistica 10 (StatSoft, Cracow, Poland).

Notably, the results of this study are presented according to hand dominance and not Parkinson’s disease dominance. First, this allowed us to directly compare patient and control participants’ results on preparatory suppression, the latter population being the one mainly studied in that domain. However, we additionally ran all analysis according to Parkinson’s dominance in the patient group only, without it appearing as a discriminative variable of interest, as validated by Principal Component Analysis (PCA) [81], which consistently selected hand dominance over Parkinson’s dominance.

## Data availability

The data supporting the findings of the present study are available upon reasonable request.

## Results

### Demographic and clinical data

A summary of demographic and clinical characteristics for each group is provided in Table 1. As expected, Parkinson’s disease patients and HC subjects did not differ in age (*P* = 0.37), but were also comparable when considering scores at the trait impulsivity questionnaire (no main effect of GROUP for the MANOVA run on the four subscales of the UPPS; *λ*_*4,52*_ = 0.85; *P* = 0.06). In contrast, but as expected, patients displayed higher depression scores at the BDI-II (*P* < 0.001).

When only looking at Parkinson’s disease patients, they scored significantly higher on the MDS-UPDRS part III when OFF-DOPA compared to ON-DOPA (*P* < 0.001), confirming the stimulation of their dopaminergic system through medication. Furthermore, as shown in Figure 2.A, the OFF-DOPA scores strongly correlated with the years since diagnosis (*R*^*2*^ = 0.30, *P* < 0.01), which ranged from 1-16 years in the current study (mean duration since diagnosis: 6.6 ± 4.2 years) (Table 2): the longer the disease duration, the higher the scores. This positive correlation was also evident when only considering the subscores for bradykinesia (*R*^*2*^ = 0.15, *P* = 0.04) (Fig. 2.A). This implies that, in our cohort, patients diagnosed a longer time ago presented a higher motor impairment [82, 83], as assessed by the MDS-UPDRS part III [58, 84]. Hence, the variable “years since diagnosis” could potentially be used as a marker of clinical progression. To verify the relevance of such an approach, in face of the variability inherent to the heterogeneity of Parkinson’s disease [85], we further clarified the evolution of clinical scores with the years since diagnosis. To do so, we performed a non-parametric Locally Estimated Scatterplot Smoothing (LOESS) regression, to assess possible non-linear relationships between both variables (Fig. 2.B), which revealed a sharp increase of OFF-DOPA clinical scores after five years since diagnosis (which appeared surrounded by two plateaus). Taken together, both the mean of the years since diagnosis as well as the visual profile of clinical scores observed with the LOESS regression in our patient cohort “agreed” on a split around six years since diagnosis. We thus split Parkinson’s disease subjects into two subgroups: one in which they had been diagnosed < 6.6 years before participating in our experiment (hereafter referred to as “PD-EARLY_STAGE_”; n=17 patients; 10 females; mean duration since diagnosis: 3.6 ± 1.5 years) and another one in which they had been diagnosed > 6.6 years before (“PD-LATE_STAGE_”; n=12 patients; 6 females; mean duration since diagnosis: 10.8 ± 2.7 years). In order to statistically investigate demographic and clinical differences between both subgroups (HC subjects were not included here), we also performed a Mann-Whitney U-test for age, BDI-II, years since diagnosis, Levodopa equivalent daily dose, MDS-UPDRS-III ON-DOPA & OFF-DOPA as well as a MANOVA for UPPS scores. As shown in Supplementary Table 2, PD-EARLY_STAGE_ and PD-LATE_STAGE_ only differed in their degree of motor symptoms, both in the ON- and OFF-DOPA state (*P* = 0.01 and *P* < 0.01, respectively) and, as expected, in their years since diagnosis (*P* < 0.001). Importantly, the mean age was comparable in both subgroups (*P* = 0.37).

**Table 2.** Demographic and clinical data in Parkinson’s disease patients and HC subjects.

|  | PD patients (n=29) |  | HC subjects (n= 29) |  | PD patients vs. HC subjects |
| --- | --- | --- | --- | --- | --- |
| | Mean (± SD) | Range (min – max) | Mean (± SD) | Range (min – max) | P-values<sup>a</sup> |
| **Age (years)** | 64.7 (± 10.5) | 41 – 79 | 62.9 (± 9.5) | 41 – 79 | $P = 0.37$ |
| **UPPS** | 87.8 (± 22.9) | 57 – 115 | 87.6 (± 15.6) | 59 – 130 | $P = 0.06$ |
| **BDI-II** | 11.8 (± 6.6) | 3 – 24 | 3.8 (± 4.8) | 0 – 21 | **$P < 0.001$** |
| **Years since diagnosis** | 6.6 (± 4.2) | 1 – 16 | - | - | - |
| **LEDD (mg)** | 700.8 (± 318.9) | 126 – 1895 | - | - | - |
| **MDS-UPDRS-III, ON-DOPA** | 22.4 (± 9.4) | 3 – 46 | - | - | - |
| **MDS-UPDRS-III, OFF-DOPA** | 30.2 (± 10.3) | 9 – 51 | - | - | - |

PD = Parkinson's disease; UPPS = UPPS Impulsive Behaviour Scale; BDI-II = Beck Depression Inventory, Second Edition; ON-DOPA = session performed one hour after the intake of dopamine replacement therapy; OFF-DOPA = session performed after overnight withdrawal of dopamine replacement therapy (min. 12 hours withdrawal for Levodopa and min. 24h withdrawal for dopamine agonists and other Parkinson's disease medication); SD = Standard Deviation.
<sup>a</sup> = P-values for PD patients vs. HC subjects comparisons (obtained from the Mann-Whitney U-test, except for the UPPS, which was analysed using a MANOVA).

**Figure 2.**
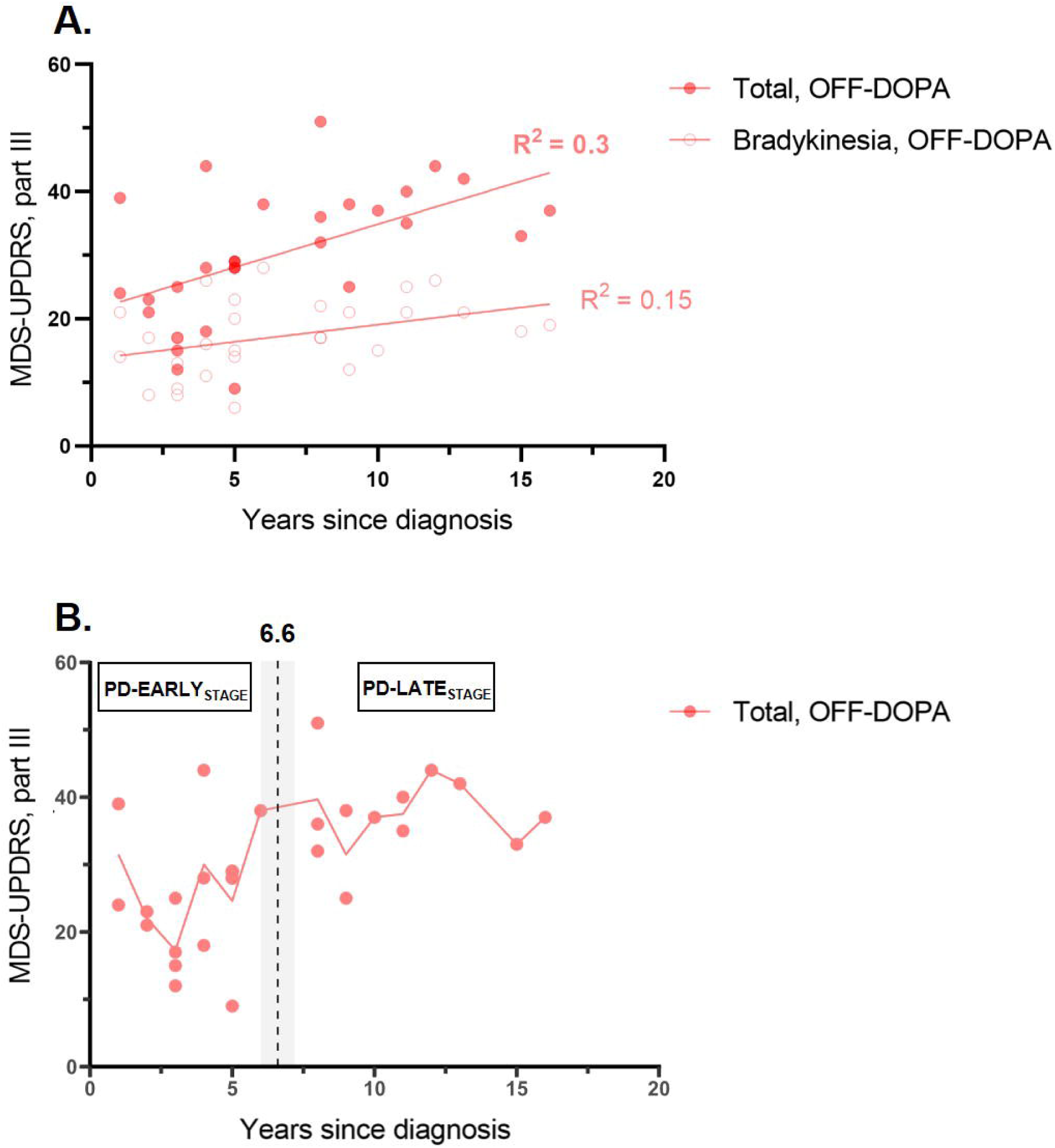
Relationship between clinical status and years since diagnosis of Parkinson’s disease. **A. OLS regressions.** A positive correlation was observed between the clinical scores and the years since diagnosis of Parkinson’s disease in the OFF-DOPA state. This was true both when considering the total scores at the MDS-UPDRS part III as well as when only considering items 3.2, 3.4, 3.5, 3.6, 3.7, 3.8 and 3.14, in order to establish a subscore for bradykinesia (*R*^*2*^ = 0.30 and *R*^*2*^ = 0.15, respectively; *P* < 0.01 and *P* < 0.05, respectively). **B. LOESS regression**. In order to further investigate the dynamics of evolution of clinical scores according to the years since diagnosis, an additional LOESS regression (span = 0.4) was performed, which revealed a pattern of increase of motor symptoms beyond five years since diagnosis, with two apparent plateaus left and right to the increase. Both types of regressions thus showed that years since diagnosis represented a suitable marker for the clinical progression of motor impairment in the disease, especially OFF-DOPA. This led us to split the patients into two subgroups, according to whether they had been diagnosed for a period shorter or longer than 6.6 years, i.e. the mean duration since diagnosis (dashed line with shadow) (PD-EARLY_STAGE_ and PD-LATE_STAGE_, respectively).

### Preparatory suppression in Parkinson’s disease patients OFF- /ON-DOPA and HC subjects

To evaluate preparatory suppression, we expressed MEP_ND_ and MEP_D_ elicited at TMS_DELAY_ in percentage of MEPs recorded at TMS_BASELINE-IN_ and analysed these percentage data using a four-way ANOVA. Interestingly, the latter revealed a significant effect of the factor GROUP [*F*(1,56) = 4.58, *P* = 0.04] due to larger percentage MEP values at TMS_DELAY_ in Parkinson’s disease patients (97.1 ± 51.3 %) than in HC subjects (81.7 ± 24.3 %), indicating overall less preparatory suppression in the patient group. Besides, analyses also yielded a significant GROUP x RESP_SIDE_ x MEP_SIDE_ interaction [*F*(1,56) = 18.65, *P* < 0.001].

As shown in Figure 3.A, healthy participants exhibited similar levels of MEP suppression during action preparation no matter the hand considered for MEPs (MEP_ND_ or MEP_D_) or the hand required for the upcoming movement response (RESP_ND_ or RESP_D_; all *P* > 0.12). Accordingly, *t*-tests run against 100 (corresponding to MEPs at TMS_BASELINE-IN_) were all significant (all *t*(28) < − 3.31 and all *P* < 0.01), confirming the systematic presence of preparatory suppression, as repetitively shown in past studies [19, 22, 26, 27]. In contrast, post-hoc analyses revealed that, in Parkinson’s disease patients, preparatory suppression varied depending on the condition: in fact, as shown in Figure 3.A, corticospinal suppression was systematically weaker when MEP_ND_ and MEP_D_ were on the side of the forthcoming movement (i.e. RESP_ND_ and RESP_D_ conditions, respectively) compared to when these MEP_ND_ and MEP_D_ were on the non-responding side (i.e. in RESP_D_ and RESP_ND_, respectively) (both *P* < 0.001). *T*-tests further supported a lack of preparatory suppression on the responding side (both *t*(28) > 0.92 and both *P* > 0.23), with percentage MEPs even leaning visually towards a facilitation. MEPs were suppressed on the non-responding side, though this effect only reached significance for MEP_ND_ (*t*(28) = − 4.94 and *P* < 0.001), but not for MEP_D_ (*t*(28) = − 1.56 and *P* = 0.13). Consistent with these findings in Parkinson’s disease patients, when comparing the level of suppression between groups, a significant difference emerged on the responding side (*P* < 0.01 and P = 0.04, for MEP_ND_ and MEP_D_ respectively), but not on the non-responding one (both *P* > 0.67).

**Figure 3.**
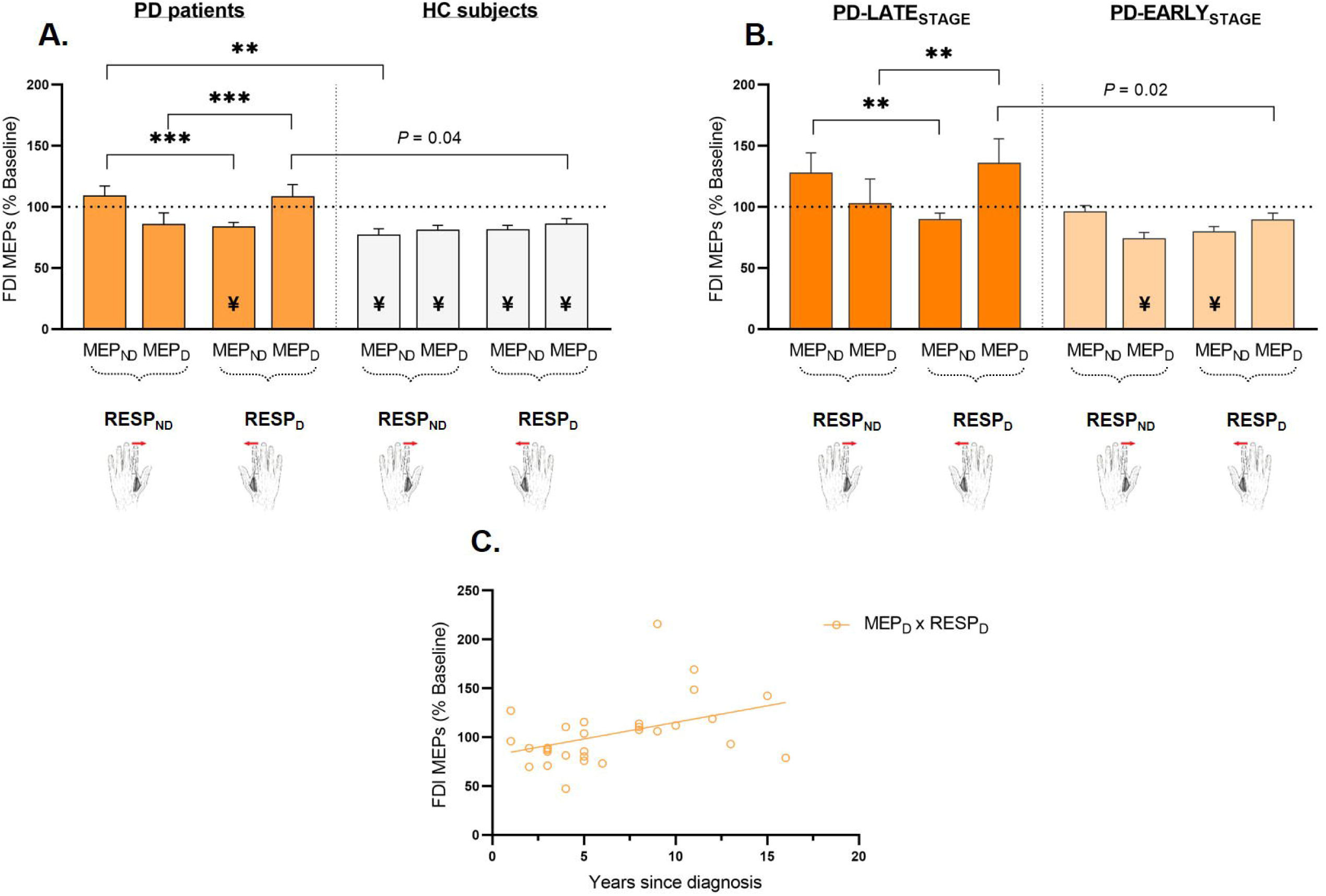
Neural measures of preparatory suppression. Amplitude of MEPs recorded at TMS_DELAY_, expressed in percentage of MEPs elicited at TMS_BASELINE-IN_, shown for the non-dominant (MEP_ND_) and dominant (MEP_D_) FDI. The hand figures represent the responding side (RESP_SIDE_) for the upcoming movement; since 93% of participants were right-handed, for the purpose of illustration, the left and right hands are used to represent non-dominant and dominant hand responses (RESP_ND_ and RESP_D_, respectively). For each group/subgroup, the bars thus represent MEP_ND_ and MEP_D_ for both FDI when they were either responding (two peripheral bars) or not responding (two central bars). Note that for every participant, the results of both sessions were pooled together, as the analysis did not reveal a significant difference between the ON- and OFF-DOPA state. **A. Comparison between Parkinson’s disease (PD) patients and HC subjects** confirmed the presence of preparatory suppression in all conditions in healthy controls (light grey), but revealed a lack of this mechanism in Parkinson’s disease (orange), in particular when the hand was selected for the forthcoming movement (peripheral bars). **B. Comparison between PD-LATE**_**STAGE**_ **and PD-EARLY**_**STAGE**_ **subgroups** showed that this lack of preparatory suppression in Parkinson’s disease mainly stemmed from patients at later disease stages (dark orange). **C. OLS regressions run between levels of preparatory suppression and years since diagnosis (in patients only)** showed a significant relationship for MEPs in the dominant hand (MEP_D_) when the latter was selected for the upcoming movement response (RESP_D_) (*R*^*2*^ = 0.18, *P* < 0.05). This indicated a progressive decline of preparatory suppression in the dominant hand with disease advancement, no matter the medication status. *T*-test, ¥ = MEPs probed at TMS_DELAY_ significantly different from those probed at TMS_BASELINE-IN_; ANOVA, **P < 0.01 and ***P < 0.001.

Critically, we did not detect any effect of medication on preparatory suppression in Parkinson’s disease patients, as revealed by the absence of a significant GROUP x DRT interaction [*F*(1,56) = 0.63, *P* = 0.43] or of any other interaction involving the factor DRT (all *P* > 0.52) (see Supplementary Fig.1 for an illustration of MEP suppression in patients according to their DRT session).

Taken together, our results indicate the existence of a deficit of preparatory suppression in Parkinson’s disease, which was most prominent on the side of the responding hand and which wasn’t restored towards physiological levels by the addition of DRT. Such findings persisted when removing a patient displaying outlier values (see Supplementary Fig. 1.A and 1.B); hence, we decided to keep this subject included for the forthcoming analysis of Parkinson’s disease subgroups.

### Preparatory suppression OFF-/ON-DOPA in PD-EARLY_STAGE_ and PD-LATE_STAGE_

Next, we addressed the question as to whether the most predominant lack of corticospinal suppression could be found at later stages of Parkinson’s disease, characterised by higher motor impairment, as illustrated by increasing clinical scores at the MDS-UPDRS part III (Fig. 2.A). To do so, we considered the TMS_DELAY_ data from the two patient subgroups in a four-way ANOVA. This analysis revealed a significant effect of the main factor SUBGROUP [*F*(1,27) = 5.92, *P* = 0.02], with less preparatory suppression observed in patients at a late compared to an early stage (Fig. 3.B). There was no significant effect of DRT [*F*(1,27) = 0.33, *P* = 0.57] or SUBGROUP x DRT interaction [*F*(1,27) = 0.89, *P* = 0.36]. Again, we found a significant SUBGROUP x RESP_SIDE_ x MEP_SIDE_ interaction [*F*(1,27) = 4.22, *P* = 0.0497].

For PD-EARLY_STAGE_, post-hoc analyses revealed comparable values, both for MEP_ND_ and MEP_D_, whether they were elicited on the side of the upcoming movement or not (both *P* > 0.07) (Fig. 3.B). The *t*-tests, however, indicated that even at an early disease stage, patients already displayed a lack of corticospinal suppression in the responding conditions (MEP_D_ in RESP_D_ and MEP_ND_ in RESP_ND_ conditions; both *t*(16) > - 1.95, both *P* > 0.07), whereas MEP suppression appeared to be preserved on the non-responding side (MEP_D_ in RESP_ND_ and MEP_ND_ in RESP_D_ conditions; both *t*(16) < - 4.97, both *P* < 0.001). Then, at later disease stages, the deficit appeared to generalize to all conditions, with PD-LATE_STAGE_ patients displaying a lack of preparatory suppression both on the responding and non-responding sides (all *t(11)* > - 2.01, all *P* > 0.07). Moreover, in these patients, the gap seemed to have grown between the two sides, with post-hoc analyses revealing significantly larger MEPs on the responding side than on the non-responding side, whether considering MEP_ND_ (larger preceding RESP_ND_ than RESP_D_, *P* < 0.01) or MEP_D_ (larger preceding RESP_D_ than RESP_ND_, *P* < 0.01) (Fig. 3.B). They also revealed that preparatory suppression differed significantly between both subgroups for MEP_D_ measures in the RESP_D_ condition (*P* = 0.02). Individual data points for PD-EARLY_STAGE_ and PD-LATE_STAGE_ (according to their DOPA session) are shown in Supplementary Fig. 1.C and 1.D, respectively; these graphs further illustrate a more severe deficit and a stronger variability in preparatory suppression in patients at later disease stages. Hence, our data suggest declining preparatory suppression over the course of Parkinson’s disease, first localized to the responding side in PD-EARLY_STAGE_ patients, but then generalizing to all conditions, including the non-responding side in PD-LATE_STAGE_ patients.

As the aforementioned ANOVA did not display any effect of DRT, we pooled together the data from both sessions when running OLS regressions between years since diagnosis and preparatory suppression. As shown in Figure 3.C, the relationship between both was significant when the latter was probed in the dominant hand (MEP_D_) when it was responding in the upcoming movement (RESP_D_ condition) (*R*^*2*^ = 0.18, *P* = 0.02). Taken together, our results thus point towards a gradual loss of preparatory suppression, particularly in the dominant hand, as disease progresses.

### Reaction and movement times in Parkinson’s disease patients OFF-/ON-DOPA and HC subjects

As shown in Figure 4, the three-way ANOVA performed on RTs did not reveal a significant effect of the main factor GROUP [*F*(1,56) = 0.54, *P* = 0.46], hence showing a similar capacity to initiate a movement response after the appearance of the imperative signal in Parkinson’s disease patients (mean RT: 247.4 ± 60.5 ms) and HC subjects (mean RT: 236.5 ± 51.2 ms) in our rolling ball task. The effect of RESP_SIDE_ [*F*(1,56) = 0.53, *P* = 0.47], GROUP x RESP_SIDE_ [*F*(1,56) = 0.10, *P* = 0.76] and GROUP x DRT [*F*(1,56) = 2.0, *P* = 0.16] interactions were not statistically significant, the latter showing the absence of an effect of medication on RTs in Parkinson’s disease patients in our task.

**Figure 4.**
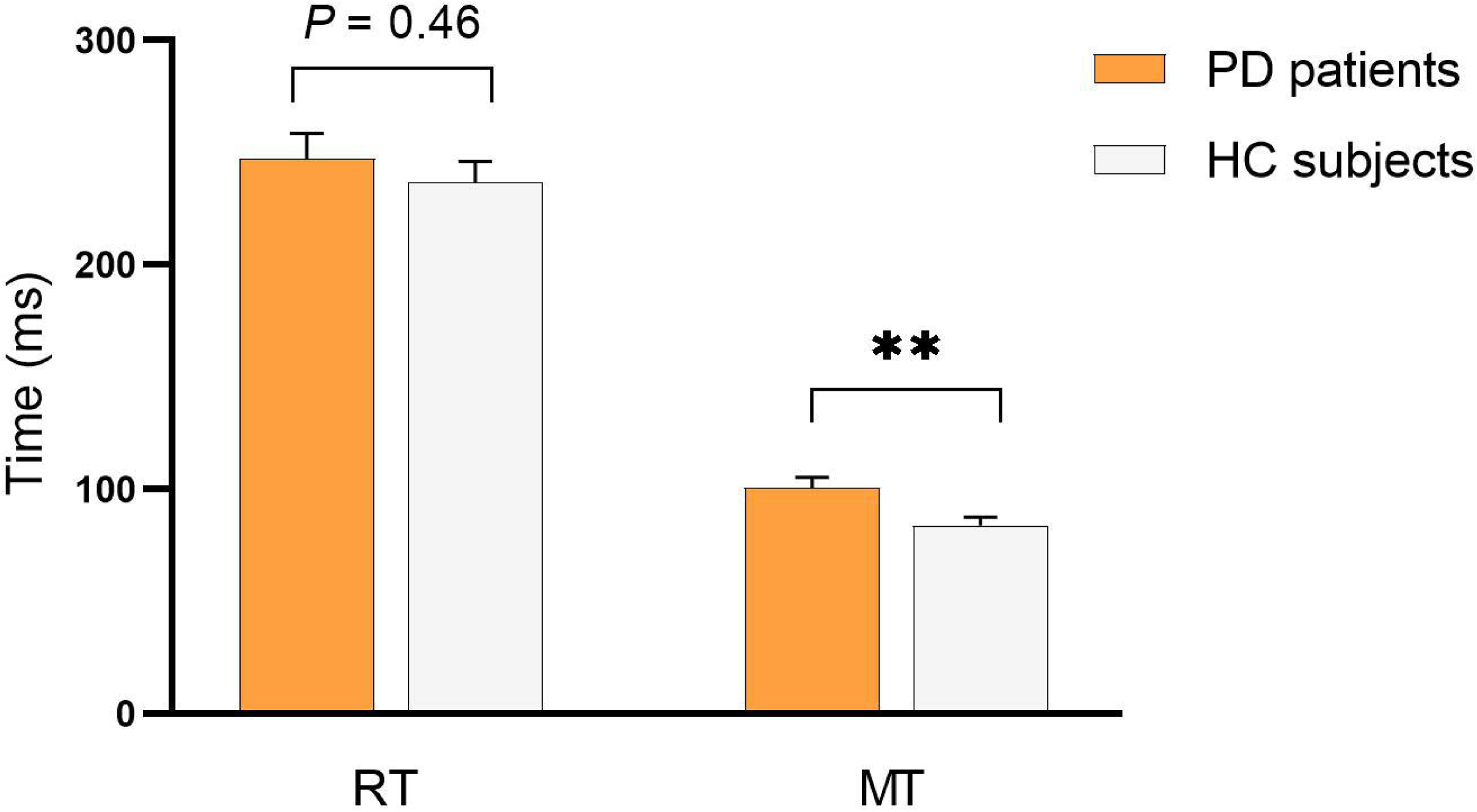
Reaction and movement times during the rolling ball task. RTs and MTs are shown for trials in which the TMS pulses were applied at rest (TMS_BASELINE-IN_) or trials in which there was no TMS pulse at all (both types pooled together) for Parkinson’s disease (PD) patients (orange) and HC subjects (light grey). RTs did not differ between both groups, but MTs were significantly longer in patients compared to control subjects. Data from both hands and both sessions are pooled together. ANOVA, **P < 0.01.

On the other hand, the same ANOVA performed on MTs yielded a significant main effect of the factor GROUP [*F*(1,56) = 7.48, *P* < 0.01], revealing longer MTs in patients (mean MT: 100.5 ± 25.8 ms) as compared to control subjects (mean MT: 83.8 ± 20.5 ms) (Fig.4). The analysis also showed a significant effect of the factor RESP_SIDE_ [*F*(1,56) = 4.23, *P* = 0.04], with longer MTs in RESP_D_ no matter the group [GROUP x RESP_SIDE_: *F*(1,56) = 0.97, *P* = 0.33]. The interaction GROUP x DRT was non-significant [*F*(1,56) = 0.42, *P* = 0.52], showing the lack of an effect of medication status on MTs.

Next, when only considering Parkinson’s disease data, no significant effect of the main factor SUBGROUP was detected for RTs [*F*(1,27) = 1.26, *P* = 0.27] or MTs [*F*(1,27) = 0.48, *P* = 0.50], showing the absence of a difference between PD-LATE_STAGE_ and PD-EARLY_STAGE_ for this type of behavioural measures, which did not appear to depend on the progression of the disease. Again, no other statistically significant interaction involving the factor SUBGROUP was observed in each respective analysis [all *F*(1,27) < 1.35, all *P* > 0.26].

Taken together, we were hence able to objectify decreased speed of movement in Parkinson’s disease patients, occurring in parallel with a less pronounced preparatory suppression. OLS regressions, however, did not allow us to proof any direct significant relationship between behavioural measures acquired during the task and measures of preparatory suppression (all *P* > 0.05).

### Raw corticospinal excitability in Parkinson’s disease patients OFF-/ON-DOPA and HC subjects Resting motor thresholds

The three-way ANOVA performed on resting motor thresholds did not reveal any significant effect of the main factor GROUP [*F*(1,56) = 0.23, *P* = 0.64] (mean: 46 ± 10% and 47 ± 9%, in Parkinson’s disease and control subjects, respectively) nor did it show a significant GROUP x DRT interaction [*F*(1,56) = 0.23, *P* = 0.63], indicating the absence of an effect of the medication status on resting motor thresholds in patients. We did not find any significant effect of the factor HEMISPHERE_SIDE_ [*F*(1,56) = 2.19, *P* = 0.15] nor a significant GROUP x HEMISPHERE_SIDE_ interaction [F(1,56) = 0.41, *P* = 0.53]. When running the three-way ANOVA on the Parkinson’s disease subgroups, no significant effect of the main factor SUBGROUP was found [*F*(1,27) = 0.02, *P* = 0.89] nor did we uncover any statistically significant interaction involving the factor SUBGROUP [all *F*(1,27) < 1.67, all *P* > 0.21].

### Baseline measures

When considering MEPs acquired at rest, at TMS_BASELINE-OUT_ and TMS_BASELINE-IN_ (i.e. outside and within the task blocks), our four-way ANOVA yielded a significant effect of the factor BASELINE [*F*(1,56) = 80.72, *P* < 0.001], without showing a significant effect of the factor GROUP [*F*(1,56) = 0.50, *P* = 0.48] nor of the GROUP x BASELINE interaction [*F*(1,56) = 1.07, *P* = 0.31]. As further revealed by post-hoc analyses, both Parkinson’s disease patients and HC subjects exhibited larger MEP amplitudes at TMS_BASELINE-IN_ (2.41 mV ±1.9 vs. 2.09 mV ±1.5, for patients and controls respectively) relative to TMS_BASELINE-OUT_ (1.42 mV ±1.3 vs. 1.30 mV ±1.2, for patients and controls respectively). This reflects an increase of corticospinal excitability within the context of the task and is in line with past research in healthy subjects [30, 86]. As shown in Figure 5, the ANOVA also yielded a significant GROUP x BASELINE x DRT x MEP_SIDE_ interaction [*F*(1,56) = 6.31, *P* = 0.02]. Post-hoc analyses revealed that, in Parkinson’s disease patients, MEPs acquired in the dominant hand (MEP_D_) were significantly larger in the OFF-DOPA as compared to the ON-DOPA state, both at TMS_BASELINE-OUT_ (*P* <0.01) and TMS_BASELINE-IN_ *(P* < 0.001). This difference between sessions in Parkinson’s disease patients was not observed for MEP_ND_, no matter the baseline condition (both *P* > 0.10). Also, when taken individually, none of the different subconditions differed between patients and controls (all *P* > 0.32), thus showing equivalent rest measures of corticospinal excitability in both groups, despite the larger MEPs OFF-DOPA compared to ON-DOPA in patients. Note that the four-way ANOVA conducted on the raw amplitude of MEPs acquired at TMS_DELAY_, i.e. while participants were preparing their movement, showed a significant GROUP x DRT x MEP_SIDE_ [*F*(1,56) = 6.89, *P* = 0.01] as well as a GROUP x RESP_SIDE_ x MEP_SIDE_ [*F*(1,56) = 25.66, *P* < 0.001] interaction, showing that the group difference in corticospinal excitability observed at baseline persisted during movement preparation. Detailed results regarding MEPs at TMS_DELAY_ are provided in the Supplementary Material.

**Figure 5.**
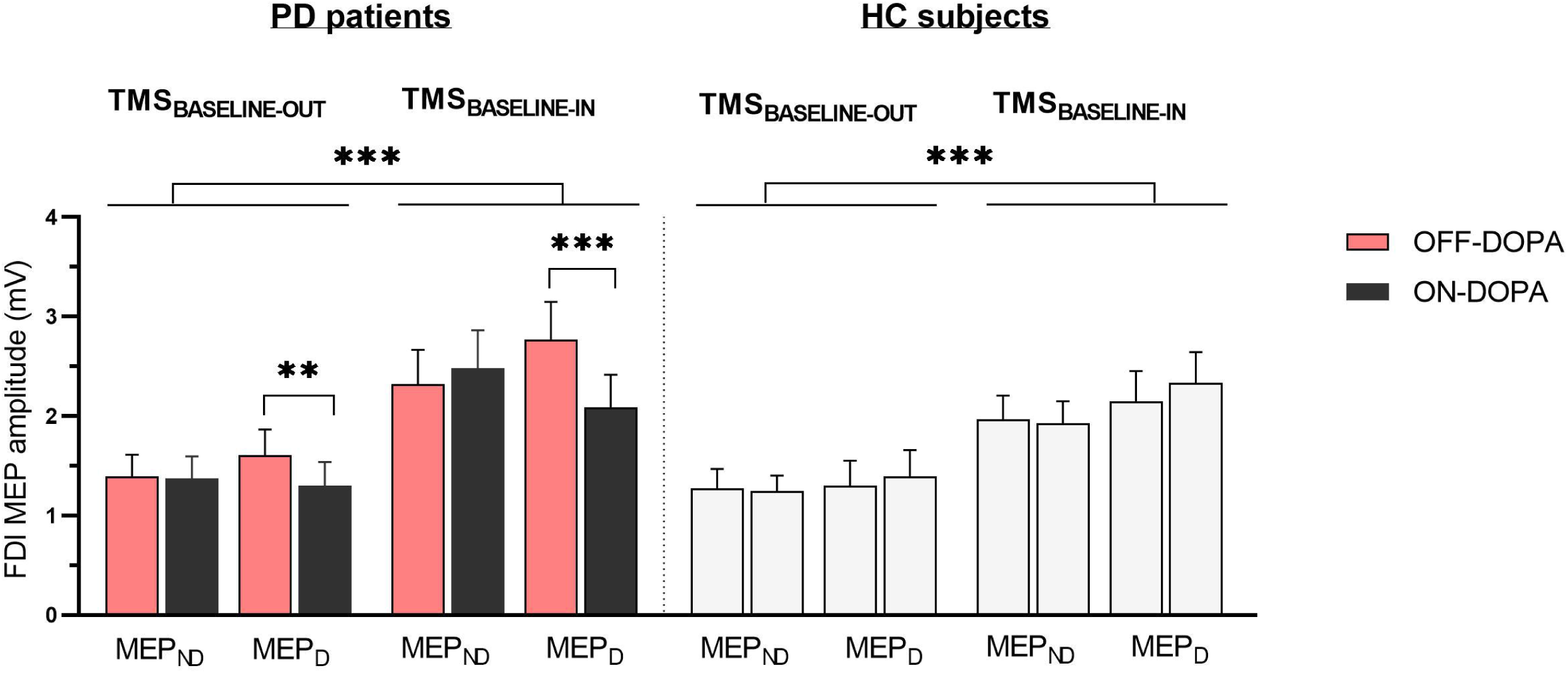
Baseline measures of corticospinal excitability. Raw amplitude of MEPs (in mV) recorded in the FDI at rest, either outside (TMS_BASELINE-OUT_) or within (TMS_BASELINE-IN_) the blocks. MEPs are shown for HC subjects (light grey) and Parkinson’s disease (PD) patients in the non-dominant (MEP_ND_) and dominant (MEP_D_) hand for the OFF-DOPA (red) and ON-DOPA (black) sessions; these sessions were fictive and randomly attributed in HC subjects. Both PD patients and HC subjects showed higher corticospinal excitability during the task as compared to complete rest. More interestingly, MEP_D_ in patients were significantly reduced in the ON-DOPA compared to the OFF-DOPA session, no matter the baseline context. ANOVA, **P < 0.01 and ***P < 0.001.

The four-way ANOVA was then repeated by considering Parkinson’s disease data only, which revealed no main effect of the factor SUBGROUP [*F*(1,27) = 0.10, *P* = 0.76] nor did it show any significant interaction involving the factor SUBGROUP [all *F*(1,27) < 2.44, all *P* > 0.13].

Taken together, we could thus show that, compared to matched healthy subjects, Parkinson’s disease patients displayed a deficit in preparatory suppression, particularly in the hand selected for the upcoming movement response, in parallel with a decreased speed of movement execution. These deficits did not seem to depend on the use of oral DRT. Next, we could confirm a most predominant lack of preparatory suppression in patients at later disease stages, who concurrently displayed an increased motor impairment as measured by the MDS-UPDRS part III. Hence, the years since diagnosis of Parkinson’s disease turned out to be a marker of the progression not only of motor symptoms OFF-DOPA, but also of the loss of preparatory suppression over time, and this for the dominant hand in particular. Finally, we were able to show that, in the same hand, raw measures of corticospinal excitability, both at rest and during movement preparation, decreased under the influence of oral dopamine intake.

### Principal component analysis

We used Principal Component Analysis (PCA) to further explore the possibility of a link between our neurophysiological (lack of preparatory suppression) and behavioural (prolonged movement times) data. We ranked variables according to their importance in the variance of the dataset to uncover potential links between them, possibly missed by OLS regressions, and also included raw neurophysiological measures. Indeed, preparatory suppression constitutes a ratio and thus encompasses the variability inherent to both raw measures of corticospinal excitability. Details of the analysis can be found in the Supplementary Material. Among our main findings here is that, no matter the PCA protocol that was run (all data vs. Parkinson’s disease data only, ON-vs. OFF-DOPA, ND vs. D), the order of the variables remained coherent, in that the analysis consistently ranked both neurophysiological as well as behavioural data first. Furthermore, links between MDS-UPDRS part III scores and raw neurophysiological measures were uncovered, which lead us to investigate if the latter might have a potential non-linear relationship with the years since diagnosis, using additional LOESS regressions.

### LOESS regressions between raw corticospinal excitability and years since diagnosis of Parkinson’s disease

In order to explore the possibility of a non-linear relationship existing between raw neurophysiological values and Parkinson’s disease progression, we decided to represent such data using LOESS regressions. As can be seen in Figure 6, showing the evolution with the years since diagnosis of raw MEP amplitudes for TMS_BASELINE-OUT_, TMS_BASELINE-IN_ and TMS_DELAY_, both in the ON- and OFF-DOPA session for both MEP_ND_ and MEP_D_, a triphasic pattern emerged: a sharp increase of corticospinal excitability after two years, dropping again towards six years (Stage 1); a plateau phase in the non-dominant hand between six and 12 years, less pronounced in the dominant hand (Stage 2); beyond 12 years, the appearance of another, less sharp, increase quickly appearing to reach another plateau (Stage 3). This recurrent, triphasic pattern indeed suggests the existence of rather dynamic changes of raw corticospinal excitability with Parkinson’s disease progression.

**Figure 6.**
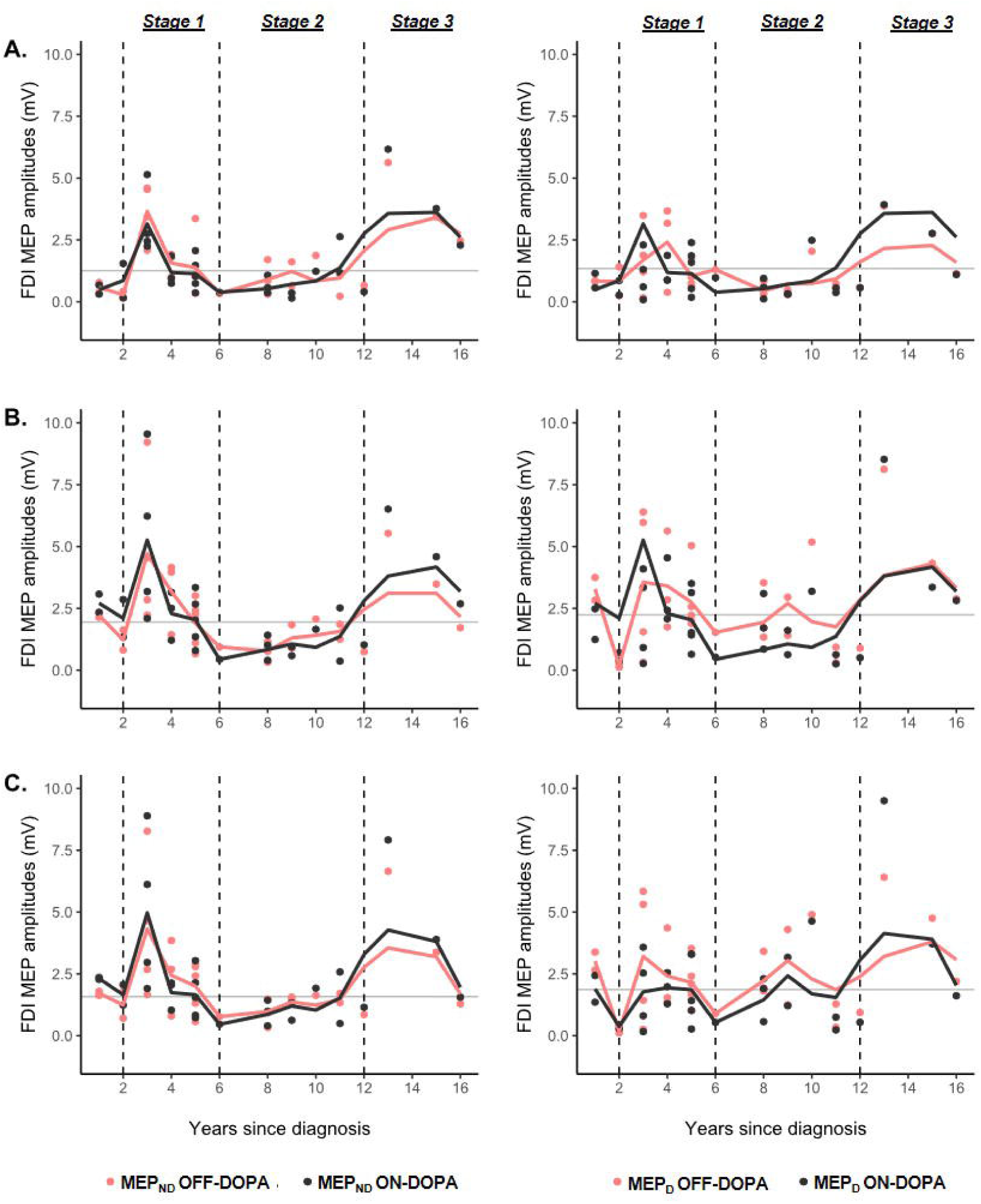
LOESS regressions illustrating the relationship between raw corticospinal excitability and years since diagnosis of Parkinson’s disease. All x-axes represent the number of years since diagnosis. Red curves and circles represent the OFF-DOPA session, while black curves and circles represent the ON-DOPA session. The light grey horizontal lines represent the mean of HC subjects in each subcondition. The three left-side graphs represent the results for the non-dominant FDI (MEP_ND_, left in most subjects) while the right-side ones represent the results for the dominant one (MEP_D_, right in most subjects). The span for all LOESS regressions was set to 0.4. **A. and B. Baseline measures**. The y-axis represents the raw amplitude of MEPs (in mV) recorded in the FDI at rest, outside (TMS_BASELINE-OUT_) and within (TMS_BASELINE-IN_) the context of the task, for A. and B., respectively. **C. Delay measures**. The y-axis represents the raw amplitude of MEPs (in mV) recorded in the FDI during the task, while participants were preparing their movement response (TMS_DELAY_). Based on the absence of an effect of RESP_SIDE_ in the significant triple interaction including DRT, MEPs from both RESP_ND_ and RESP_D_ conditions were pooled together for each FDI. As highlighted by the vertical dashed lines, a triphasic pattern emerged for all three TMS timings, which likely represents alterations of raw corticospinal excitability due to different adaptive and compensatory mechanisms over the course of Parkinson’s disease (Stage 1 – 3).

To the best of our knowledge, there exists no longitudinal data on the progression of alterations of raw corticospinal excitability in Parkinson’s disease. Hence, in the face of our sample size, in order to further challenge the patterns observed in our LOESS regressions, we proceeded to synthetic data amplification using CTGAN, a machine learning algorithm to model tabular data. Details regarding this method are presented in the Supplementary Material. Supplementary Fig. 4 illustrates how the patterns observed in our cohort held up when data was amplified by 100%, thus doubling the numbers of data points. This means that the patterns of evolution of raw neurophysiological measures with disease progression observed in our cohort have a high probability of surviving in bigger patient populations.

## Discussion

In order to further elucidate how abnormal functional changes within M1 in Parkinson’s disease are linked to cardinal motor symptoms like bradykinesia, we propose a novel approach specifically investigating patterns of changes in corticospinal excitability underlying voluntary movement preparation. We found that patients exhibited a lack of corticospinal suppression during action preparation, as compared to what is typically observed in healthy subjects, no matter their medication status. Interestingly, this deficit was most predominant in patients at later compared to earlier stages of the disease as well as in the dominant hand. We also found that raw MEP amplitudes in patients were globally larger OFF-compared to ON-DOPA when elicited in the dominant hand. Concomitantly, we detected an increasing motor handicap with disease progression as well as objective motor slowness during the task in our patient cohort. Even if such alterations did not correlate directly with neurophysiological measures, our findings are in line with the idea that a lack of preparatory suppression may disrupt fast movement execution by altering motor neural gain.

### Preparatory suppression fades in late Parkinson’s disease independently of dopamine medication

Our findings in healthy subjects are in line with studies reporting a strong suppression of corticospinal excitability during movement preparation [19, 21, 26], regardless of whether a trial requires moving the dominant or non-dominant hand and whether MEPs are obtained on the side of the responding or non-responding hand [25, 30, 72]. It is worth noting that preparatory suppression was substantial in our healthy control group, even though participants were on average markedly older than those typically included in other papers [29, 31] and there exists evidence of an age-related decline in this mechanism [87]. Yet, our findings here indicate that, even so, preparatory suppression is still notable in healthy aging [88].

Parkinson’s disease patients, however, displayed a lack of this mechanism, which was most prominent in later disease stages. In the latter case, MEPs systematically failed to show a suppression during movement preparation and corticospinal excitability even seemed to become facilitated though non-significantly on the side of the responding hand, particularly the dominant one. Contrariwise, patients in earlier disease stages exhibited a profile of corticospinal suppression that came close to the one of healthy subjects, in that it was preserved on the non-responding side; preparatory suppression did however already present signs of weakness in these patients too when considering MEPs on the responding side. Interestingly, the latter deficit was found to correlate with the disease stage: the more advanced the disease, the more patients displayed a lack of preparatory suppression in the dominant responding hand. This finding points towards a progressive loss of this motor control mechanism over the course of Parkinson’s disease − evident first (and then worsening) on the body side concerned with the upcoming movement response and affecting the non-responding side only at later disease stages in parallel with gradual dopaminergic neurodegeneration [46, 89, 90]. Importantly, the decline of preparatory suppression observed in Parkinson’s disease cannot be accounted for by a difference in age given that the latter was comparable in the two patient subgroups; rather, this finding supports the idea that it relies on operational nigro-striatal dopaminergic projections. Yet paradoxically, we did not observe any difference in the degree to which MEPs were suppressed between the OFF- and ON-DOPA sessions: that is, the lack of preparatory suppression was present even when patients were medicated, no matter the disease stage. It should be noted here that, since we found highly significant improvements in UPDRS scores, it is unlikely that missing effects were caused by an excessive residual stimulation of the dopaminergic system OFF medication [91].

Although surprising at first sight, this is in line with the work of other authors previously reporting that deficits in higher motor control are poorly responsive to DRT [92, 93]. Such mechanisms, like preparatory suppression, typically entail both purely motor as well as cognitive aspects of motor control [19, 21, 26]. Interestingly, studies in Parkinson’s disease have shown that cognitive deficits in particular are poorly responsive to dopaminergic treatment, as opposed to motor impairments [94, 95], the latter being typically used as the reference mark to adapt DRT dosage. Whereas motor deficits arise from early dopamine deficiency in the putamen leading to alterations in the “motor” fronto-striatal loop including the premotor cortex (PMC) [91, 96], cognitive dysfunction is linked to depletion of dopamine in the caudate nucleus appearing later in the disease and affecting the “cognitive” loop including the prefrontal cortex (PFC) [97]. This progressively extending striatal neurodegeneration over time, as well as the resulting differential effects of dopaminergic medication on motor and cognitive aspects of higher motor control, could explain the gradually expanding loss of preparatory suppression as disease advances, while also clarifying the absence of an effect of DRT on such a mechanism.

Interestingly, in healthy subjects, preparatory suppression in responding hands has been linked to inhibitory inputs from the PMC, whereas the lateral PFC has been shown to have inhibitory effects on both the non-responding and responding hands, thus orchestrating a more global corticospinal suppression during movement preparation [19, 98]. In Parkinson’s disease, several studies have demonstrated PMC hyperactivity with facilitatory influences onto M1, which has been interpreted as an early compensatory mechanism to help movement initiation [14, 44, 91, 99]. Such findings could explain why the responding hand conditions in our patient cohort were altered first: early PMC hyperactivity could potentially lead to an altered capacity to exert inhibitory influences while preparing a movement with a selected effector. In contrast, preparatory suppression on the non-responding side is linked to activity within the PFC, which has been shown to decrease with Parkinson’s disease progression [100], possibly linked to the spreading of neuropathology over time [101] or reflecting gradual alterations in the activity of neural networks underlying cognition [102]. Alternatively, the decline in preparatory suppression over the years might reflect the increasing involvement of other non-dopaminergic neurotransmitter systems [93], in particular the cholinergic one [103].

### Deficient preparatory suppression comes along with increased motor slowness in Parkinson’s disease

In our rolling ball task, Parkinson’s disease patients showed similar RTs but longer MTs than the control group, no matter their medication status. The RT findings are in fact consistent with past research showing that, even if simple RTs are typically prolonged in Parkinson’s participants, choice RT results can be variable [3, 8]. Our MT findings, on their side, did objectively demonstrate the presence of motor slowness in our patient cohort as compared to healthy participants [11, 15].

The fact that, in patients, slower movements occurred in parallel with a lack of preparatory suppression in responding hands is in line with the idea that a stronger suppression of motor circuits during movement preparation yields faster movement execution [22, 31]. This corroborates the gain modulation hypothesis [26, 32], whereby suppression of motor cortical background activity would increase neuronal sensitivity to excitatory drives, thus accelerating movement execution in selected effectors; the most prominent lack of preparatory suppression in our patient cohort was indeed found in responding hands, especially the dominant one. In the face of an apparent loss of preparatory suppression with the years since diagnosis of Parkinson’s disease, in parallel with increasing dopaminergic neurodegeneration [46], we propose that dopamine could be at the source of such gain modulation [38]. However, we observed no effect of DRT on patients’ task behaviour, in line with results of preparatory suppression. In Parkinson’s disease, the effects of dopaminergic medication on RTs have been variable, although choice RTs appear to be more sensitive than simple RTs [3, 10, 16]. For MTs, studies typically show an improvement of movement amplitude and velocity under dopaminergic medication [15, 45]; it should be noted, though, that such findings are often obtained outside the context of tasks probing higher motor control. This could also explain the contrast with the clinical scores obtained before the task at the MDS-UPDRS part III, which did improve under dopaminergic medication in our cohort. Finally, the absence of an effect of medication on our MT findings could also be linked to the limited abduction amplitude potentially missing slight variations between the ON- and OFF-DOPA session. The latter could also explain that unlike for clinical scores and preparatory suppression and contrary to what can be found in the literature [11] we did not objectify a decrease of movement speed with the years since diagnosis. Finally, it should be noted that we could not yet formally establish a significant correlation between MT and MEP changes. This resonates with past studies in healthy subjects, which have acknowledged the difficulty to relate the degree of MEP suppression to movement parameters, probably because the size of MEPs is dependent on several overlapping mechanisms, precluding a straightforward link with specific behavioural features [26].

### Dopamine medication globally decreases raw measures of corticospinal excitability

Despite comparable levels of preparatory suppression and movement parameters between both sessions in patients, we did observe an effect of dopamine medication on corticospinal excitability, as raw MEPs in their dominant hand were larger in the OFF-compared to the ON-DOPA state, both at rest (within and between blocks) and during the preparatory delay (no matter the responding hand). Dopaminergic medication has been typically reported to normalize M1 excitability measures [3]. However, there seems to be no clear relationship between levodopa-related improvements in movement kinematics and TMS measures, which possibly indicates that both have a different sensitivity to dopamine [3, 15, 44, 45]; this could further explain the absence of a medication effect on our behavioural measures.

The fact that only the dominant hand showed medication-related changes in corticospinal excitability points towards a hyperexcitable state of that effector in particular, when OFF-DOPA [15]. The higher use of the dominant hand in daily life might indeed have as a signature the need for stronger, global compensatory excitatory influences in order to facilitate motor readiness, especially in the absence of medication. Such compensation could possibly be orchestrated by the PMC which as mentioned above has been shown to be hyperactive early in Parkinson’s disease, particularly without DRT [91, 99]. However, of notice is that the various raw MEP measures OFF-DOPA in our patients did not significantly differ from the equivalent measures in control subjects, which can appear in contradiction with research showing increased M1 activity and corticospinal excitability in Parkinson’s disease compared to healthy subjects [12, 14]. Such findings might be partially explained by the fact that raw corticospinal excitability displayed considerable variations according to the disease stage, which could have masked the existence of group differences in our cohort (Fig.6).

### Raw measures of corticospinal excitability likely evolve with Parkinson’s disease progression

LOESS regressions run on our data to illustrate the evolution of raw MEPs according to the years since diagnosis of Parkinson’s disease allowed to detect the emergence of recurrent, triphasic patterns according to specific disease stages (Fig.6). Those persisted after data amplification with novel deep learning methods for visualisation purposes, which allows to emit hypotheses on a possible pattern of evolution of corticospinal motor output with disease progression. According to Blesa et al. (2017), once intrinsic basal ganglia compensatory mechanisms following the start of dopaminergic neurodegeneration fail, excessive inhibitory output onto the thalamo-M1 projection probably leads to the appearance of the first parkinsonian motor symptoms and motor cortical abnormalities under the form of a hyperactive and hyperexcitable M1 [12, 14, 41, 100, 104, 105], early in the disease; this is coherent with Stage 1 of our observed pattern (Fig. 6). Since motor cortical activity and corticospinal output depend on a subtle interplay between facilitatory and inhibitory inputs [19], such hyperexcitability can reflect alterations to one or the two of these inputs; both of them have been shown to be altered in Parkinson’s disease [41, 105]. This is thought to possibly constitute a compensatory mechanism, to help generate sufficient corticospinal output despite reduced inputs from basal ganglia [15, 106]. Upstream areas like the PMC are potential candidates that could exert such compensation, especially in the OFF-DOPA state [91, 99]. Over time, high cortical excitability can disrupt encoding of motor parameters [107] which could contribute to altered processing of inputs from upstream areas [15], leading to the appearance of altered functional connectivity between the PMC and M1 [44, 108]. This way, compensatory mechanisms could turn less effective, having as a consequence a decrease in corticospinal excitability, visible particularly in the non-dominant hand (Stage 2, Fig. 6); this could lead to gradually worsening motor symptoms, particularly in the OFF-DOPA state. Interestingly, the decrease in raw corticospinal excitability in Stage 2 coincided with the increase in clinical scores in our cohort (Fig. 2.B). Finally, Stage 3 (Fig. 6) was characterised by another increase in corticospinal excitability, which might be the signature of more extensive network alterations [56, 109], possibly linked to the topographical extension of neuropathology [101, 110] and the functional and structural alterations appearing in higher motor control centres such as the PFC; this could alter their capacity to exert control over M1. Taken together, increased cortico-spinal excitability, which can be beneficial in the early stages of Parkinson’s disease, can over time significantly alter correct movement implementation, among others through perturbations in capacities to exert gain modulation within the motor system. Our interpretation could reconcile two seemingly opposed hypotheses advanced by Bologna et al. (2018), stating that increased M1 excitability had to be either an adaptation mechanism or a deleterious mechanism: it could actually represent a continuous spectrum of mechanisms whose net result over time turns out detrimental for movement execution.

### Limitations and future directions

Although we used disease progression in our analyses, we here want to recognize again that “years since diagnosis of Parkinson’s disease” is not a perfect variable as there is heterogeneity inherent to a diagnosis being established by different movement disorder specialists. Furthermore, it should be noted that this disorder is characterised by a complexity to disentangle direct effects of the disease and compensatory changes that can develop over time [56]. The longitudinal nature inherent to compensatory processes renders interpretations based on cross-sectional data less robust [12]; thus, study designs that further explore questions related to M1 excitability changes in the future should ideally be of longitudinal nature.

Finally, as we had to exclude patients with significant tremor but also dyskinesia, in order to assure correct task performance, we indirectly had to disregard patients at the most advanced disease stages, typically those awaiting second-line treatments [111]. One way to include patients with very long disease durations is to recruit them after surgical treatments such as deep brain stimulation, a patient sub-population which is currently examined in a follow-up study.

Future study protocols aiming at investigating preparatory suppression in Parkinson’s disease could benefit from multimodal approaches, like combining TMS and neuroimaging or TMS-EEG [112] and/or from tasks that allow to disentangle the motor and cognitive aspects of such a motor control mechanism.

## Conclusions

Our study provides novel insights into alterations of the motor system in Parkinson’s disease, by uncovering a clear deficit in corticospinal suppression typically shaping neural activity during movement preparation in healthy populations. Our results point towards a gradual loss of this marker of intact motor control with Parkinson’s disease progression, in parallel with increasing dopaminergic neurodegeneration. This could be the signature of an alteration, in this clinical population, of dopamine-related mechanisms of motor neural gain necessary for proper movement execution, as illustrated by their decreased movement velocity and increasing motor handicap. While dopaminergic medication did not restore abnormalities linked to preparatory suppression, it nevertheless appeared to globally decrease raw corticospinal excitability in the dominant hand. Our findings thus suggest differential functional roles of dopamine in shaping corticospinal motor output in Parkinson’s disease and illustrate the importance of considering disease progression when investigating such outputs. Principal component analysis allowed to show early evidence for a relationship between neurophysiological and movement parameters, supporting the importance of moving to more task-related functional markers, such as preparatory suppression, when studying motor impairment in Parkinson’s disease.

## Data Availability

All data produced in the present study are available upon reasonable request to the authors

## Abbreviations

DRT: dopamine replacement therapy
FDI: first dorsal interosseous
HC: healthy control
M1: primary motor cortex
LOESS: locally estimated scatterplot smoothing
MDS-UPDRS: Movement Disorder Society Unified Parkinson’s Disease Rating Scale
MEP: = motor evoked potential
MT: movement time
OLS: ordinary least squares
PCA: principal component analysis
RT: reaction time
TMS: transcranial magnetic stimulation

## Acknowledgements

First, the authors wish to express their deep gratitude to all patients and control subjects who participated in the current research project, despite a complicated sanitary context being in place for a lot of them at the moment of their testings. Next, the authors would like to thank Prof. Ivanoiu, Dr. Leempoel, Dr. Peeters and Dr. Verougstraete (neurologists at the Saint-Luc University Hospital), as well as “Association Parkinson” and “Action Parkinson”, for their valuable help in the recruitment of patient participants. The authors also wish to thank Vincent Leroux and Brice Virlée for their relevant contribution to participants’ recruitment and data collection, as well as the centre for support in statistical methodology and calculation (*SMCS*) of the *UCLouvain*. This paper is dedicated to Prof. Jeanjean, the former head of the Adult Neurology department and renowned movement disorder specialist at the Saint-Luc University Hospital, as well as the former thesis co-supervisor and clinical mentor of author EW.

## Author contributions

EW, CQ, GD, AJ and JD designed the research protocol; EW and AJ recruited the participants; EW conducted the experiments; JD supervised the study; EW, CQ, GD, SP and JD analysed the anonymised data; EW wrote the manuscript; EW, CQ, GD, SP and JD reviewed the manuscript.

## Supplementary material

Supplementary material is available online.

