## Supplementary material for "Corticospinal suppression underlying intact movement preparation fades in late Parkinson’s disease"

#### Raw corticospinal excitability during movement preparation in Parkinson's disease patients OFF-/ON-DOPA and HC subjects

In addition to the analyses of preparatory suppression and of corticospinal excitability at rest, we also considered the raw amplitude of MEPs (mV) acquired at $TMS_{DELAY}$. Those raw measures were analysed by conducting a four-way ANOVA, with GROUP (Parkinson's disease patients, HC subjects) as the between-subject factor and with DRT (ON-DOPA, OFF-DOPA), $RESP_{SIDE}$ ($RESP_{ND}$, $RESP_{D}$) and $MEP_{SIDE}$ ($MEP_{ND}$, $MEP_{D}$) as within-subject factors. The four-way ANOVA conducted on the raw amplitude of MEPs acquired at $TMS_{DELAY}$, i.e. while participants were preparing their movement, first showed a significant GROUP x DRT x $MEP_{SIDE}$ interaction [$F(1,56) = 6.89$, $P = 0.01$]. Post-hoc analyses revealed that in Parkinson's disease patients, just as for the baseline measures, $MEP_{D}$ were significantly larger in the OFF-DOPA as compared to the ON-DOPA state ($P < 0.01$), no matter the $RESP_{SIDE}$ (see Supplementary Fig. 2). The ANOVA also yielded a significant triple GROUP x $RESP_{SIDE}$ x $MEP_{SIDE}$ interaction [$F(1,56) = 25.66$, $P < 0.001$]. As shown by post-hoc analyses, both the amplitude of $MEP_{ND}$ and of $MEP_{D}$ differed between the $RESP_{ND}$ and $RESP_{D}$ conditions (both $P < 0.001$), with higher MEP amplitudes being found in both FDIs when the muscle was selected for the forthcoming movement (i.e. $MEP_{ND}$ and $MEP_{D}$ in the $RESP_{ND}$ and $RESP_{D}$ conditions, respectively) than when it was not selected (i.e. $MEP_{ND}$ and $MEP_{D}$ in the $RESP_{D}$ and $RESP_{ND}$ conditions, respectively). When taken individually, none of the different subconditions differed between patients and controls (all $P > 0.09$), thus showing equivalent raw delay measures of corticospinal excitability for both groups, in concordance with the rest measures presented in the main text.

The four-way ANOVA was then repeated by considering the two Parkinson's disease subgroups only. No significant effect of the main factor SUBGROUP was found [$F(1,27) = 0.04$, $P = 0.85$] nor did any statistically significant interaction involving the factor SUBGROUP appear in the analysis [all $F(1,27) < 3.92$, all $P > 0.06$].

### Principal component analysis

Small datasets can suffer from inherent variability, raising the question as to whether results obtained in them can be extrapolated to bigger cohorts. Principal component analysis (PCA) is a well-established technique with data exploratory potential: it identifies the principal components responsible for most of the variance of the dataset and ranks them from the first (which explains most of the variance) to the last (which explains least of the variance). It then converts correlations (or lack thereof) among all variables into 2D graphs, in a "dimension reduction" fashion; variables which are highly correlated cluster together. The PCA algorithm thus increases the interpretability of the dataset while also minimizing information loss.

In the Parkinson's disease literature, PCA has so far mainly been used in the context of neuroimaging studies[1,2] or studies focusing on diagnostic and symptom monitoring tools[3-5]. To the best of our knowledge, it hasn't been used yet in the context of neurophysiological studies focusing on patterns of corticospinal excitability and of related motor behaviour or symptoms in Parkinson's disease patients.

Here, the standard protocol for PCA was applied with the function *PCA* from the *R* package "FactoMineR"; the *fviz_pca* visualizations from the package "factoextra" were used to guide the development of the analyses and configure the illustrations[6]. The *corrplot* function (https://github.com/taiyun/corrplot) from the package "corrplot" was used to generate a standardized correlation matrix of the variables according to the principal components.

The PCA protocol was run on both the whole data set (Parkinson's disease patients and HC subjects) as well as on patients only. For both of them, we first considered all the data, then we ran PCA according to the hand dominance (ND vs. D only) or according to the session (ON-DOPA vs. OFF-DOPA only). We wanted to evaluate if certain variables would show higher correlational profiles with regard to certain (sub)groups (Parkinson's disease patients vs. HC subjects, PD-EARLY$_{STAGE}$ vs. PD-LATE$_{STAGE}$). As discussed in the main text, when only considering the patient group, we also ran PCA according to Parkinson's disease dominance, in order to analyse its potential for being a discriminative variable, as compared to hand dominance. As "years since diagnosis" could become a major distinctive variable between patients, it was removed from all PCA. With the iterations of the PCA development, the variables with minor importance to variance and those not relevant to the question in check were progressively removed in order to clarify the inner relationships between the variables that remained.

In the PCA performed for both the whole dataset as well as for patients only, we successively accounted for the ON- vs. OFF-DOPA division and the difference in hand dominance. Taken together, results were homogeneous, as the raw neurophysiological variables ($TMS_{BASELINE-IN}$ and $TMS_{DELAY}$) consistently came to explain most of the variance of the dataset, appearing either in the first (PC1) or second principal component (PC2). In the PCA run on patients only, both total and bradykinesia scores at the MDS-UPDRS-III typically dominated PC1, reinforcing the validity of the clinical tests. Furthermore, we also persistently saw MT and RT appear with correlation potential to preparatory suppression, especially OFF-DOPA. In the face of these relatively consistent results across all protocols run, we here present the PCA performed on patients only, comparing their ON- and OFF-DOPA session, as it highlights the aforementioned findings and paves the way for interesting correlations to be further explored in the future.

First, in order to assess the validity of the PCA, we generated a cumulative variance plot (Supplementary Fig. 3.A), showing that by dimension 4, we had reached 75% of explicability for the variance of the dataset, which assured us that this PCA had a high informative value and that we could proceed with its use. Next, the matrix in Supplementary Fig. 3.B displays correlations on a scale from dark blue (strong negative correlation) to dark red (strong positive correlation). PC1 and PC2 appear similar, highlighting the importance of both clinical scores and raw MEP amplitudes. As shown on the biplot of Supplementary Fig. 3.C, representing PC1 (x-axis) and PC2 (y-axis), clinical scores (both ON- and OFF-DOPA) appeared to co-evolve with OFF medication values of preparatory suppression and MTs, as well as RTs (both ON- and OFF-DOPA), all being positively correlated with PC1 (quadrant 1). Along the positive side of PC2, in quadrant 2, MT ON medication seemed to co-evolve with raw MEP amplitudes at $TMS_{BASELINE-IN}$ and $TMS_{DELAY}$ (both ON- and OFF-DOPA). Interestingly, along PC1 and PC2, MDS-UPDRS part III scores and raw neurophysiological values evolved in opposite directions, which lead us to investigate if the latter might have a potential non-linear relationship with disease progression, using additional LOESS regressions (Fig. 6, main text).

Taken together, our PCA findings support further explorations in the future of the relationship between neurophysiological, behavioural and clinical variables in bigger Parkinson's disease cohorts, in order to mitigate the variability inherent to smaller datasets.

#### Modelling tabular data using conditional GAN

Modelling the probability distribution of rows in tabular data and generating realistic synthetic data is a non-trivial task: tabular data usually contains a mix of discrete and continuous columns. Continuous columns may have multiple modes whereas discrete columns are sometimes imbalanced making the modelling difficult. Existing statistical and deep neural network models fail to properly model this type of data. Conditional tabular generative adversarial network (CTGAN) was designed to address these challenges. CTGAN has been shown to outperform Bayesian methods on most of the real datasets whereas other deep learning methods don't (https://arxiv.org/abs/1907.00503). It has thus become the gold standard for synthetic data amplification techniques.

We used CTGAN (package: https://github.com/sdv-dev/CTGAN; function: CTGANSynthesizer) to amplify our Parkinson's disease data for raw MEP amplitudes at $TMS_{BASELINE-OUT}$, $TMS_{BASELINE-IN}$ and $TMS_{DELAY}$. We used 10000 epochs (number of iterations in the learning process) and a K-Fold cross validation as learning method. Next, we ran LOESS regressions with the years since diagnosis. We did this to challenge the question as to whether our observed patterns would hold up in bigger patient populations, since to the best of our knowledge there exist no longitudinal data on the progression of alterations of raw corticospinal excitability in Parkinson's disease. As show in Supplementary Fig. 4, with an amplified dataset of 60 rows (31 new samples, for a total of 60 patient data points), the patterns observed in our cohort persisted, allowing us to infer that they have a high probability to remain and become more defined in larger patient populations.

### Supplementary tables

**<u>Supplementary Table 1.</u> Average number of trials remaining per condition after data cleaning in Parkinson's disease patients and HC subjects.**

| | **PD patients** | | | | **HC subjects** | | | |
|---|---|---|---|---|---|---|---|---|
| | **$MEP_{ND}$** | | **$MEP_D$** | | **$MEP_{ND}$** | | **$MEP_D$** | |
| **$TMS_{BASELINE\text{-}OUT}$** | 42.1 ± 3.2 | | 42.2 ± 2.9 | | 42.8 ± 3.1 | | 42.8 ± 2.4 | |
| **$TMS_{BASELINE\text{-}IN}$** | 26.8 ± 5.9 | | 26.5 ± 6.6 | | 29.0 ± 5.3 | | 29.0 ± 4.8 | |
| **$TMS_{DELAY}$** | $RESP_{ND}$ | $RESP_D$ | $RESP_{ND}$ | $RESP_D$ | $RESP_{ND}$ | $RESP_D$ | $RESP_{ND}$ | $RESP_D$ |
| | 26.4 ± 7.4 | 26.2 ± 7.3 | 26.4 ± 7.2 | 25.0 ± 7.5 | 30.4 ± 5.5 | 31.0 ± 5.1 | 30.5 ± 5.1 | 30.2 ± 5.4 |

PD = Parkinson's disease; $TMS_{BASELINE\text{-}OUT}$ = pulse delivered outside the context of the task, at complete rest in front of a blank screen; $TMS_{BASELINE\text{-}IN}$ = pulse delivered during the inter-trial interval; $TMS_{DELAY}$ = pulse delivered during the preparatory delay, between the appearance of the preparatory cue and the imperative signal; $MEP_{ND}$ and $MEP_D$ = motor evoked potentials probed in the non-dominant and dominant hand, respectively; $RESP_{ND}$ and $RESP_D$ = non-dominant and dominant hand selected for providing the upcoming movement response, respectively.

**<u>Supplementary Table 2.</u> Demographic and clinical data in the PD-EARLY$_{STAGE}$ and PD-LATE$_{STAGE}$ patient subgroups.**

| | **PD-EARLY$_{STAGE}$ (*n*=17)** | | **PD-LATE$_{STAGE}$ (*n*=12)** | | **PD-EARLY$_{STAGE}$ vs. PD-LATE$_{STAGE}$** |
|---|---|---|---|---|---|
| | **Mean (± SD)** | **Range (min – max)** | **Mean (± SD)** | **Range (min – max)** | ***P*-values [a]** |
| **Age (years)** | 62.7 (± 11.9) | 41 – 79 | 67.4 (± 7.9) | 55 – 79 | $P = 0.38$ |
| **UPPS** | 86.8 (± 17.2) | 57 – 114 | 89.4 (± 29.9) | 59 – 115 | $P = 0.72$ |
| **BDI-II** | 9.8 (± 5.7) | 3 – 21 | 14.7 (± 7.2) | 7 – 24 | $P = 0.08$ |
| **Years since diagnosis** | 3.6 (± 1.5) | 1 – 6 | 10.8 (± 2.7) | 8 – 16 | **$P < 0.001$** |
| **LEDD (mg)** | 671 (± 398) | 126 – 1895 | 743.1 (± 158.8) | 460 – 953 | $P = 0.23$ |
| **MDS-UPDRS-III, ON-DOPA** | 19.2 (± 10.1) | 3 – 46 | 26.9 (± 6.1) | 16 – 37 | **$P = 0.01$** |
| **MDS-UPDRS-III, OFF-DOPA** | 25.1 (± 9.5) | 9 – 44 | 37.5 (± 6.5) | 25 – 51 | **$P < 0.01$** |

PD = Parkinson's disease; BDI-II = Beck Depression Inventory, Second Edition; UPPS = UPPS Impulsive Behaviour Scale; ON-DOPA = session performed one hour after the intake of dopamine replacement therapy; OFF-DOPA = session performed after overnight withdrawal of dopamine replacement therapy (min.12 hours withdrawal for Levodopa and min. 24h withdrawal for dopamine agonists and other Parkinson's disease medication); SD = Standard Deviation.

[a] = *P*-values for PD-EARLY$_{STAGE}$ vs. PD-LATE$_{STAGE}$ comparisons (obtained from the Mann-Whitney U-test, except for the UPPS, which was analysed using a MANOVA).

### Supplementary figure legends

<u>Supplementary Figure 1.</u> **Neural measures of preparatory suppression in Parkinson's disease (PD) patients only (individual data).** Amplitude of MEPs recorded at $TMS_{DELAY}$, expressed in percentage of MEPs elicited at $TMS_{BASELINE\text{-}IN}$, shown for the non-dominant ($MEP_{ND}$) and dominant ($MEP_{D}$) FDI in each patient participant, according to the OFF-DOPA (red) and ON-DOPA (black) session. The hand figures represent $RESP_{SIDE}$ for the upcoming movement; since 86% of the patients were right-handed, for the purpose of illustration, the left and right hands are used to represent non-dominant and dominant hand responses ($RESP_{ND}$ and $RESP_{D}$, respectively). For each session, the bars thus represent $MEP_{ND}$ and $MEP_{D}$ for both FDI when they were either responding (two peripheral bars) or not responding (two central bars). **A. Individual data points, excluding outlier subject 19** (S19). **B. Individual data points, including outlier subject 19**, who exhibited a considerable lack of preparatory suppression, especially in the ON-DOPA session. Both figures confirm the global tendency for a lack of preparatory suppression in Parkinson's disease patients, especially in the selected hand conditions, no matter the medication status. **C. and D. show the individual data points of PD-EARLY$_{STAGE}$ and PD-LATE$_{STAGE}$, respectively**, illustrating a more marked lack of preparatory suppression and more variability in patients at later stages of the disease.

<u>Supplementary Figure 2.</u> **Raw corticospinal excitability at $TMS_{DELAY}$.** Raw amplitude of MEPs (in mV) recorded in the FDI during the task, while participants were preparing their movement response ($TMS_{DELAY}$). MEPs are shown for Parkinson's disease (PD) patients (red and black) and HC subjects (light grey) in the non-dominant ($MEP_{ND}$) and dominant ($MEP_{D}$) hand. Based on the results of the ANOVA, $RESP_{ND}$ and $RESP_{D}$ measures are pooled together. Red bars represent the OFF-DOPA whereas black bars represent the ON-DOPA session; such sessions were fictive and randomly attributed in HC subjects. $MEP_{D}$ in patients were significantly reduced in the ON-DOPA session compared to the OFF-DOPA session, no matter the $RESP_{SIDE}$. ANOVA, $**P < 0.01$.

<u>Supplementary Figure 3.</u> **Principal component analysis (PCA) for the Parkinson's disease patients group only. A. Cumulative variance plot.** The x-axis shows the principal components, while the y-axis illustrates the cumulative variance explained by them. The plot shows that >75% of the variance (dashed line) of the dataset was explained within the first four dimensions of principal components, which proofed that a PCA was a valid method to explore

the data included in this study. **B. Correlation matrix.** The matrix showed the importance, within the first five principal components (PC1-PC5), of the different variables included in the PCA. Dark blue represents strong negative correlations, while dark red represents strong positive correlations; the more intense the color, the more significant the correlation. **C. Biplot.** Represented in this biplot are the different variables projected along the first and second principal components (PC1 on the x-axis, PC2 on the y-axis). Along PC1, clinical scores seemed to co-evolve with values of preparatory suppression and MT, especially OFF-DOPA, as well as RTs. Along PC2, MTs ON-DOPA appeared to co-evolve with raw MEP amplitudes at $TMS_{BASELINE\text{-}IN}$ and $TMS_{DELAY}$. Taken together, this PCA highlights the importance of further investigating the relationship between neurophysiological, behavioural and clinical motor variables in bigger Parkinson's disease cohorts.

**Supplementary Figure 4.** **LOESS regressions illustrating the relationship between raw corticospinal excitability and years since diagnosis of Parkinson's disease, after synthetic data amplification using CTGAN.** All x-axes represent the number of years since diagnosis. Red curves and circles represent the OFF-DOPA session, while blue curves and circles represent the ON-DOPA session. The light grey horizontal lines represent the mean of HC subjects in each subcondition. The three left-side graphs represent the results for the non-dominant FDI ($MEP_{ND}$, left in most subjects) while the right-side ones represent the results for the dominant one ($MEP_{D}$, right in most subjects). The span for all LOESS regressions was set to 0.4. Data was amplified by ~100% (31 additional samples), hence bringing the number of data points to a total of 60 as compared to our original dataset. **A. and B. Baseline measures.** The y-axis represents the raw amplitude of MEPs (in mV) recorded in the FDI at rest, outside ($TMS_{BASELINE\text{-}OUT}$) and within ($TMS_{BASELINE\text{-}IN}$) the context of the task, for A. and B., respectively. **C. Delay measures.** The y-axis represents the raw amplitude of MEPs (in mV) recorded in the FDI during the task, while participants were preparing their movement response ($TMS_{DELAY}$). Based on the absence of an effect of the $RESP_{SIDE}$ in the triple interaction including DRT, MEPs from both $RESP_{ND}$ and $RESP_{D}$ conditions were pooled together for each FDI. These LOESS regressions run on synthetically amplified data showed that the triphasic patterns observed in our original data set held up when data points were increased with machine learning algorithms, using CTGAN. Such patterns thus have a high probability of surviving in larger Parkinson's disease patient cohorts.

### Supplementary figures

#### Supplementary Figure 1.

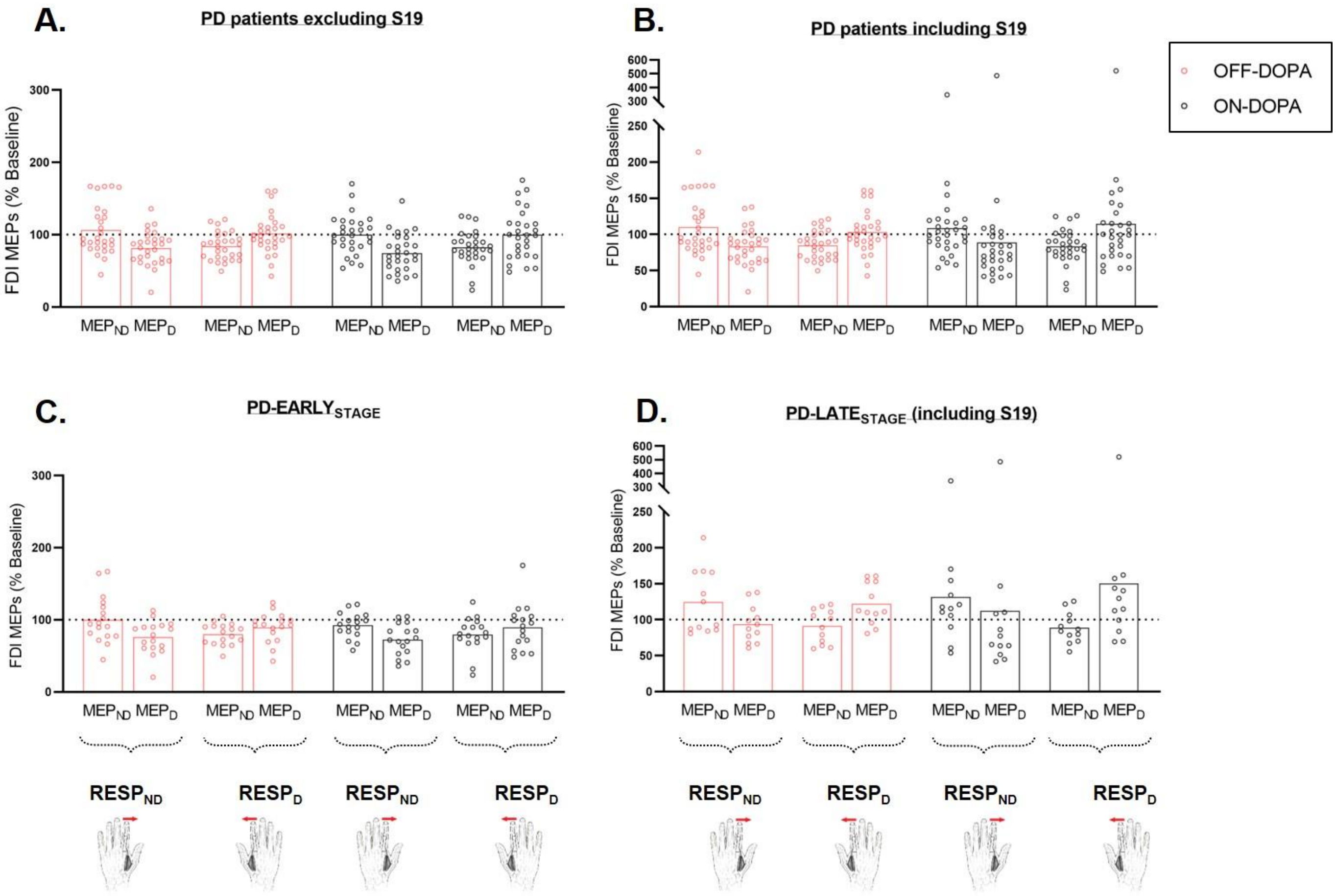


#### Supplementary Figure 2.

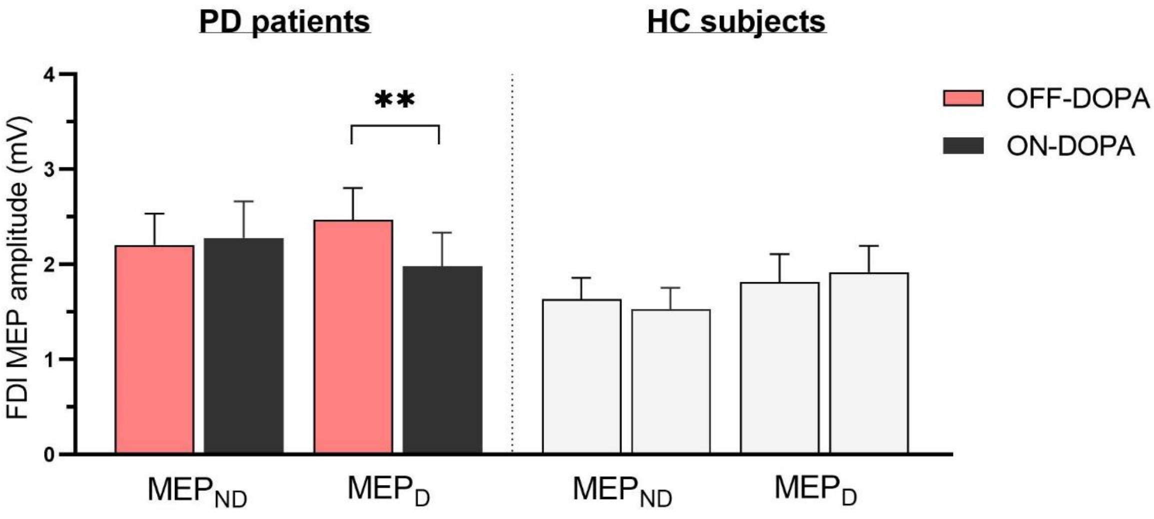

**Supplementary Figure 3.**

A.

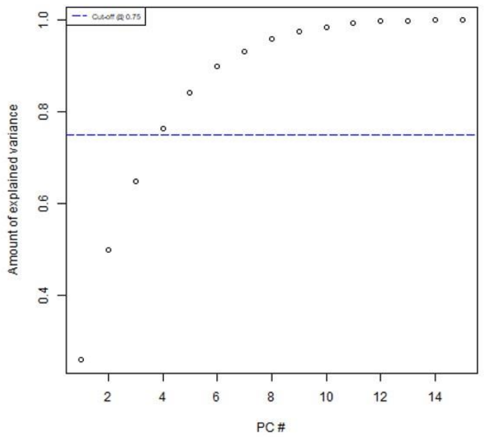


B.

| | PC1 | PC2 | PC3 | PC4 | PC5 |
|---|---|---|---|---|---|
| MT ON-DOPA | -0.06 | -0.07 | 0.34 | -0.48 | 0.22 |
| RT ON-DOPA | 0.24 | -0.13 | -0.48 | 0.06 | 0.32 |
| MT OFF-DOPA | 0.13 | -0.12 | 0.19 | -0.25 | 0.66 |
| RT OFF-DOPA | 0.19 | -0.02 | -0.45 | 0.24 | 0.34 |
| LEDD | -0.09 | -0.08 | 0.33 | -0.04 | 0.27 |
| MDS-UPDRS-III ON-DOPA | 0.29 | -0.38 | 0.08 | -0.03 | -0.18 |
| MDS-UPDRS-III OFF-DOPA | 0.31 | -0.35 | 0.02 | 0.01 | -0.24 |
| Bradykinesia ON-DOPA | 0.31 | -0.34 | 0.18 | 0.05 | -0.03 |
| Bradykinesia OFF-DOPA | 0.32 | -0.32 | 0.15 | 0.10 | -0.11 |
| TMS(BASELINE-IN) ON-DOPA | -0.35 | -0.34 | -0.06 | 0.10 | 0.05 |
| TMS(DELAY) ON-DOPA | -0.33 | -0.35 | -0.16 | -0.06 | -0.05 |
| Preparatory Suppression ON-DOPA | 0.00 | 0.02 | -0.24 | -0.59 | -0.35 |
| TMS(BASELINE-IN) OFF-DOPA | -0.36 | -0.33 | -0.05 | 0.07 | 0.04 |
| TMS(DELAY)OFF-DOPA | -0.34 | -0.35 | -0.15 | -0.05 | 0.04 |
| Preparatory Suppression OFF-DOPA | 0.14 | -0.03 | -0.37 | -0.52 | 0.06 |

C.

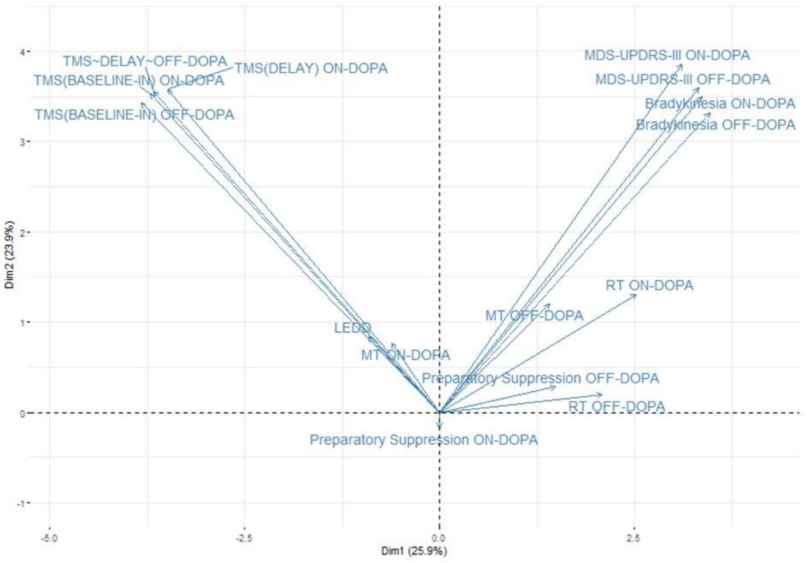

**Supplementary Figure 4.**

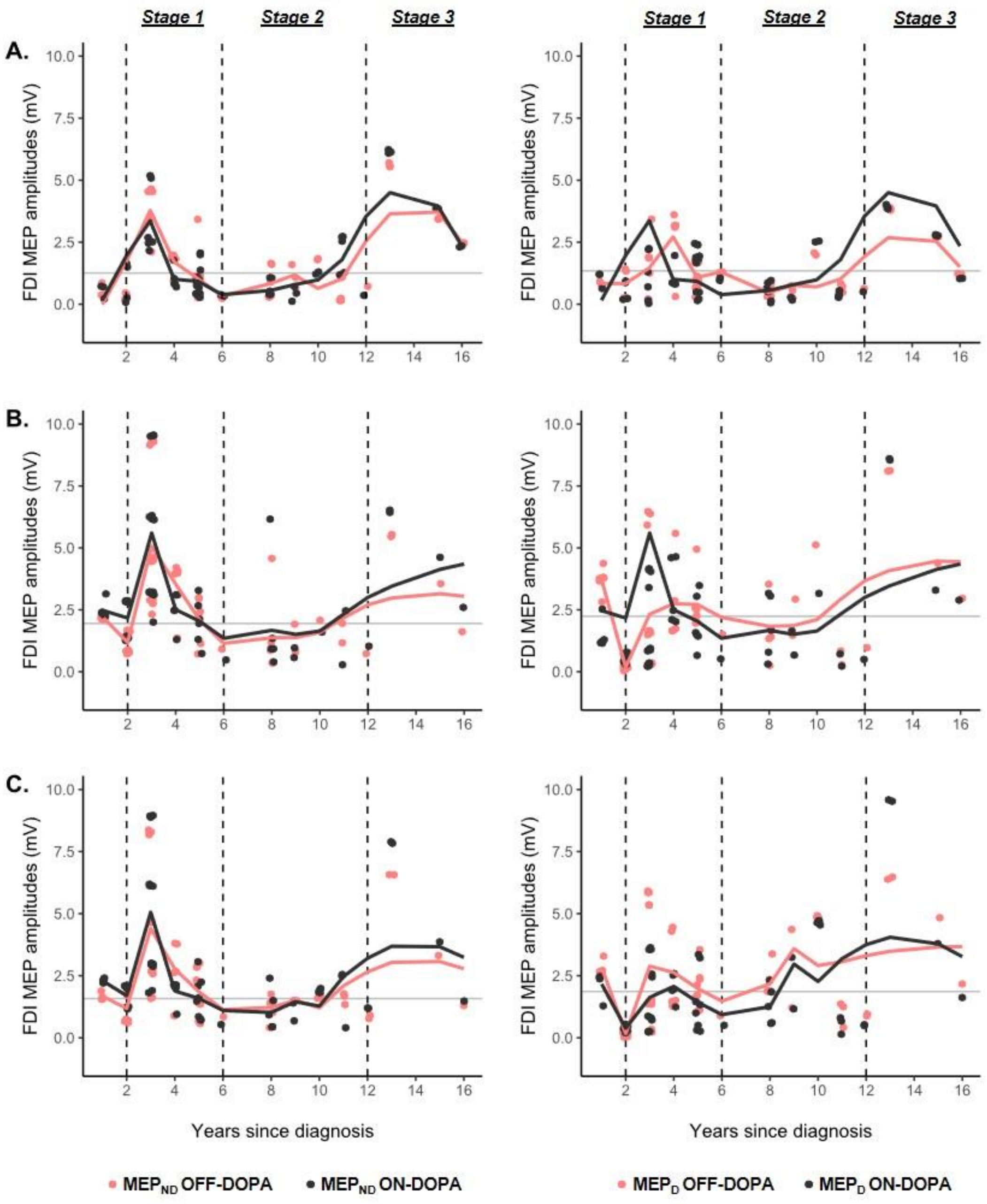
